# Effect of herpes zoster vaccination on dementia occurrence: A quasi-experimental study in Australia

**DOI:** 10.1101/2024.06.27.24309563

**Authors:** Michael Pomirchy, Christian Bommer, Fabienne Pradella, Felix Michalik, Ruth Peters, Pascal Geldsetzer

**Affiliations:** Division of Primary Care and Population Health, Department of Medicine, Stanford University, Stanford, California, USA; Heidelberg Institute of Global Health, Heidelberg University, Heidelberg, Germany; Gutenberg School of Management and Economics, Mainz University, Mainz, Germany; Ageing and Neurodegeneration, Neuroscience Research Australia, Sydney, Australia; School of Psychology, University of New South Wales, Sydney, Australia; Ageing Futures Institute, University of New South Wales, Sydney, Australia; Neurology, The George Institute for Global Health, Sydney, Australia; The Phil and Penny Knight Initiative for Brain Resilience at the Wu Tsai Neurosciences Institute, Stanford University, Stanford, California, USA; Department of Epidemiology and Population Health, Stanford University, Stanford, California, USA; Chan Zuckerberg Biohub – San Francisco, San Francisco, California, USA

## Abstract

**Importance:** Taking advantage of a natural experiment in Wales, our group has recently provided evidence that herpes zoster (HZ) vaccination appears to prevent or delay dementia. Exploiting a similar natural experiment in Australia, this present study investigated the effect of HZ vaccination on dementia occurrence in a different population and health system setting.

**Objective:** To determine the effect of HZ vaccination on the probability of receiving a new diagnosis of dementia in the future.

**Design, Setting, and Participants:** In Australia, starting on November 1 2016, live-attenuated HZ vaccination was provided for free to individuals aged 70 to 79 years of age through primary care providers. Thus, those whose 80^th^ birthday was just a few weeks prior to November 1 2016 never became eligible, whereas those whose 80^th^ birthday was just a few weeks later were eligible. The key strength of our approach is that one would not expect that these comparison groups who differ in their age by only a minute degree would, on average, differ in any of their health characteristics and behaviors. We analyzed primary healthcare records with week-of-birth information from 65 general practices across Australia using regression discontinuity.

**Exposure:** Eligibility for HZ vaccination based on one’s date of birth.

**Main outcome:** New diagnoses of dementia as recorded in primary care electronic health record data.

**Results:** As expected, in our sample of 101,219 patients, individuals born just before versus shortly after the date-of-birth eligibility threshold (November 2 1936) for HZ vaccination were well-balanced in their past preventive health services uptake and chronic disease diagnoses. There was an abrupt increase of 16.4 (95% CI: [13.2 – 19.5], p < 0.001) percentage points in the probability of ever receiving HZ vaccination between patients born shortly before versus shortly after the date-of-birth eligibility threshold. The eligibility rules of the HZ vaccination program, thus, created comparison groups just on either side of the date-of-birth eligibility threshold who were likely similar to each other, except for a large difference in their probability of receiving the intervention (HZ vaccination) of interest. Drawing on a sample of 18,402 patients, we find that eligibility for HZ vaccination (i.e., being born shortly after versus shortly before November 2 1936) decreased the probability of receiving a new dementia diagnosis over 7.4 years by 1.8 percentage points (95% CI: [0.4 – 3.3], p = 0.013). Being eligible for HZ vaccination did not affect the probability of taking up other preventive health services (including other vaccinations), nor the probability of being diagnosed with other common chronic conditions than dementia.

**Conclusions and Relevance:** Corroborating our quasi-experimental findings from Wales in a different population, this study provides important evidence on the potential benefits of HZ vaccination for dementia because its quasi-experimental design allows for conclusions that are more likely to be causal than those of more commonly conducted associational studies.

**Key points:** *Question:* What is the effect of herpes zoster vaccination on the probability of receiving a new diagnosis of dementia in the future?

*Findings:* In this quasi-experimental study of electronic health record data from Australia, being eligible for herpes zoster vaccination based solely on one’s date of birth significantly decreased the probability of receiving a new dementia diagnosis over 7.4 years by 1.8 percentage points. **Meaning:** Due to its quasi-experimental design, this study provides evidence for a beneficial effect of herpes zoster vaccination for preventing or delaying dementia that is more likely to be causal than the associations reported in the existing correlational evidence.

## Introduction

Neurotropic herpesviruses have long been thought to potentially play a causative role in the development of dementia.^1–5^ Herpes zoster (HZ) vaccination may, therefore, have a protective effect on the development of dementia. A second reason for which HZ vaccination could have benefits for dementia is that there is evidence, especially in the case of live-attenuated vaccines, that vaccines have important off-target health effects induced by broader immune mechanisms.^6–8^

Our group has recently shown in Wales-wide electronic health record data that HZ vaccination significantly reduced the probability of receiving a new dementia diagnosis over the subsequent seven years.^9^ By taking advantage of a natural experiment, our study in Wales overcame the fundamental limitation of the existing, exclusively associational,^10–20^ evidence on HZ vaccination and dementia that those who opt to be vaccinated differ from those who do not in a variety of characteristics that are difficult to measure and that could, thus, confound the findings.^21^ For instance, detailed information on health behaviors that are likely related to both dementia and vaccination, such as physical activity and diet,^22,23^ are virtually never available in electronic health record data.

This study exploits a similar natural experiment as in Wales to investigate the effect of HZ vaccination on the occurrence of dementia in a different population and health system setting. Specifically, we take advantage of the fact that only those aged 70 to 79 years on November 1 2016, when the Australian National Immunisation Programme (NIP) started its HZ vaccination program, were eligible for free live-attenuated HZ vaccination (Zostavax [Merck]).^24–26^ Thus, individuals who had their 80^th^ birthday just prior to, or on, November 1 2016 (i.e., born before November 2 1936) were ineligible for HZ vaccination whereas those who had their 80^th^ birthday just after November 1 2016 were eligible. This eligibility rule resulted in an abrupt increase in the probability of ever receiving the HZ vaccine between individuals who differed in their age by merely a week across the date of birth-based eligibility threshold for the vaccination program. By comparing these groups just on either side of the date of birth-based eligibility threshold, the Australian setting, thus, allows for a comparison of dementia incidence between eligible and ineligible groups of individuals who are not expected to differ in their characteristics (including health behaviors for which information is not available in electronic health record data) other than a minute difference in age and a large difference in the probability of ever receiving the HZ vaccine.

## Methods

### The herpes zoster vaccine rollout in Australia

Australia’s National Immunisation Programme (NIP), first introduced in 1997, is a collaborative program between the Australian State and Territory governments that provides free vaccines to eligible individuals with the goal of preventing diseases.^27^ The NIP for HZ vaccination started on November 1 2016.^24^ As of this date, the live-attenuated single-dose HZ vaccine (Zostavax [Merck]) was provided free of charge nationwide in Australian primary care practices for those aged 70 to 79 years. Thus, individuals born on or after November 2 1936 (i.e., those who had their 80^th^ birthday after November 1 2016) were eligible for free HZ vaccination, whereas those born before November 2 1936 (i.e., those who had their 80^th^ birthday before or on November 1 2016) were ineligible and remained ineligible for life. Further information on the HZ vaccination rollout in Australia is available elsewhere.^24–26^

### Data source

We used data from PenCS,^28^ an Australian-owned health informatics company, which provides detailed primary care electronic health records to researchers. The data included diagnoses, immunizations and other healthcare procedures, as well as prescribed medications from 65 general practitioner (GP) practices across each of Australia’s six states and the Australian Capital Territory. More detail on these practices is available in **Text S1**.

For the purposes of our analysis, PenCS provided us with patients’ dates of birth in weeks. As is customary in Australia’s primary care records, diagnoses were coded by PenCS using open-ended text fields provided by the GP. The text fields used to define each diagnosis in our analysis are listed in **Table S1**. PenCS does not link any of its primary care records to hospital records or mortality registers.

### Selection/exclusion criteria for patients

Our analysis was limited to patients who were at least 50 years of age on November 1 2016 and who have visited one of 65 GP practices in Australia between February 15 1993 and March 27 2024. Figure 1 presents a flowchart describing the cohort development and our selection/exclusion criteria for our primary analyses.

**Fig. 1:**
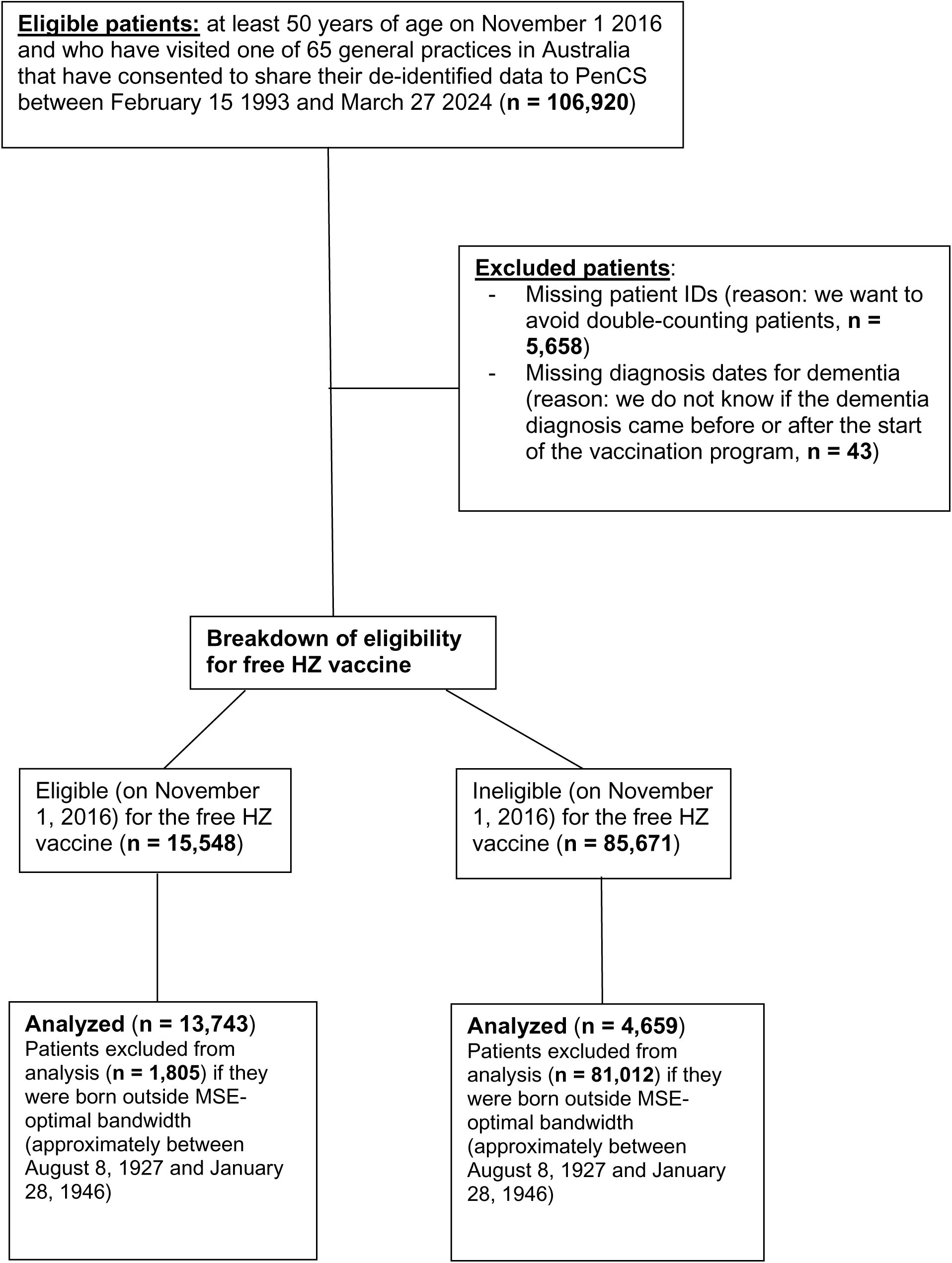
**Flowchart of Development of PenCS Patient Sample** Abbreviations: HZ = Herpes Zoster; MSE = Mean Squared Error

### Outcome and exposure definitions

Our follow-up period began on the start date (November 1 2016) of the HZ vaccination program. Our dataset ended in March 27 2024, which marked the end of the follow-up period.

Our outcome of interest was new diagnoses of dementia made during the follow-up period. If more than one diagnosis for dementia was recorded for an individual patient, we used the date of the first diagnosis. This approach of using the date of the first diagnosis was also used for defining the date of all other diagnoses in our analyses. Given the neuropathological overlap between dementia types and the difficulty in distinguishing dementia types clinically,^29–31^ as well as our reduced statistical power when studying less common outcomes, we defined dementia as dementia of any type or cause. The codes used to define dementia (as well as all other diagnoses used in our analyses) are listed in Table S1.

Our exposure was eligibility for free HZ vaccination as determined by an individual’s date of birth. Week of birth in our data was coded such that each week started on a Monday. Because November 2 1936 was also a Monday, we were thus able to determine the eligibility status of each patient in the data.

### Statistical analysis

We used three methodological approaches in this paper, which are described in more detail (along with all robustness checks and tests for confounding) in **Text S2**. First, we used a regression discontinuity (RD) design, which exploits the discontinuity in eligibility for a free HZ vaccination at the date-of-birth eligibility threshold (i.e., November 2 1936). This analysis is based on the rationale that individuals born very close to either side of the November 2 1936 threshold are expected to be similar to each other in observed and unobserved characteristics except for their eligibility status for HZ vaccination. In addition to restricting the analysis to a bandwidth around the threshold, the RD design, thus, assigns the highest weights (by using triangular kernel weighting) to those individuals born most closely around the November 2 1936 threshold. The key strength of RD is that it provides an unbiased effect estimate as long as any confounding variables do not abruptly change at exactly the November 2 1936 date-of-birth threshold.^32,33^

The RD design provides unbiased effect estimates even in the presence of censoring as long as the degree of censoring does not abruptly change between individuals at the November 2 1936 date-of-birth threshold. Nonetheless, in secondary analyses, we accounted for the different amounts of follow-up time across patients by modeling dementia as a time-to-event outcome.

Our first time-to-event approach was a cause-specific accelerated failure time (AFT) model, using the approach by Adeleke et al.,^34^ who have specifically adapted this model to RD settings. In our second time-to-event approach, we used an alternative approach to RD, termed the local randomization approach,^32,35^ to determine a narrow bandwidth around the November 2 1936 eligibility threshold in which patients can be expected to be exchangeable with each other. For the sample of patients within this narrow bandwidth, we then created Kaplan-Meier plots for each treatment condition and compared cumulative incidence curves between eligible and ineligible patients using Gray’s test.

Finally, we used a variant of the RD design, called comparative RD (CRD), in secondary analyses, by leveraging an additional source of untreated data from older patients in our sample. By adding these data, CRD tends to provide increased statistical power relative to standard RD.^36^ In addition to the main cohort of patients who were born in close proximity to the date of birth-based eligibility threshold, our CRD design analyzed a comparison cohort of patients that was always ineligible for free HZ vaccination.

### Ethics

This research was approved by the Stanford University Institutional Review Board on June 9 2023 and considered minimal risk (protocol number: 70277).

## Results

### Characteristics of the study population

Our dataset contained data on 101,219 unique patients. **Table 1** shows the sociodemographic and clinical characteristics of this sample. We additionally show the characteristics of the 18,402 patients born within the mean squared error (MSE)-optimal bandwidth (for our primary analysis on the effect of HZ vaccination on new diagnoses of dementia) of 482 weeks around the November 2 1936 date-of-birth eligibility threshold.

**Table 1.** Characteristics of the PenCS sample and eligible and ineligible patients at the date-of-birth eligibility threshold.^1,2,3,4,5^. Abbreviations: MSE = Mean Squared Error, TIA = Transient Ischemic Attack; PPV = Pneumococcal Polysaccharide Vaccine; DPT = Diphtheria, Tetanus, and Pertussis ^1^ The MSE-optimal bandwidth used in our primary analysis for the effect of HZ vaccination on new diagnoses of dementia was 482 weeks. ^2^ 1,620 (1.6%) patients in the entire sample and 234 (1.3%) patients in the MSE-optimal bandwidth had missing information on gender. ^3^ The clinical diagnoses shown are dementia and the 15 most common diagnoses in our data. The codes used to define each condition are shown in Table S1. All diagnoses are defined as being recorded prior to November 1 2016. ^4^ The codes used to define each indicator of preventive health services uptake are shown in Table S1. All indicators were defined as being recorded prior to November 1 2016. ^5^ Cancer screening refers to the uptake of colorectal or breast cancer screening. As per Australian cancer screening guidelines, this was defined as uptake of fecal occult blood testing (for colorectal cancer screening) and mammography (for breast cancer screening).^56,57^

|  | Sample within<br>MSE-optimal<br>bandwidth | Eligible<br>patients at the<br>threshold | Ineligible<br>patients at the<br>threshold | Difference in<br>means at the<br>threshold (p-value) |
| --- | --- | --- | --- | --- |
| ***Sociodemographic characteristics*** |  |  |  |  |
| Female | 9,992 (55.0%) | 53.5% | 55.8% | -2.3% (p = 0.188) |
| Married | 5,402 (29.4%) | 28.5% | 26.6% | 1.9% (p = 0.209) |
| Aboriginal or Torres<br>Strait Islander | 128 (0.7%) | 0.7% | 0.9% | -0.1% (p = 0.689) |
| Foreign-born | 455 (2.5%) | 2.7% | 2.5% | 0.2% (p = 0.712) |
| ***Clinical diagnoses*** |  |  |  |  |
| Hypertension | 2,928 (15.9%) | 18.1% | 17.8% | 0.4% (p = 0.787) |
| Hyperlipidemia | 1,948 (10.6%) | 11.5% | 11.9% | -0.4% (p = 0.688) |
| Respiratory<br>conditions | 1,291 (7.0%) | 6.5% | 7.9% | -1.4% (p = 0.113) |
| Osteoarthritis | 1,539 (8.4%) | 9.7% | 9.9% | -0.2% (p = 0.835) |
| Non-hematological<br>cancers | 1,442 (7.8%) | 10.1% | 9.3% | 0.8% (p = 0.423) |
| Heart conditions | 1,660 (9.0%) | 10.8% | 10.7% | 0.1% (p = 0.902) |
| Diabetes mellitus | 1,120 (6.1%) | 6.2% | 5.6% | 0.6% (p = 0.485) |
| Depression | 472 (2.6%) | 2.2% | 2.2% | -0.1% (p = 0.896) |
| Osteoporosis | 871 (4.7%) | 5.2% | 6.5% | -1.3% (p = 0.110) |
| Back pain | 357 (1.9%) | 2.6% | 1.8% | 0.8% (p = 0.103) |
| Gout | 444 (2.4%) | 2.7% | 2.9% | -0.2% (p = 0.719) |
| Stroke or TIA | 447 (2.4%) | 2.8% | 2.7% | 0.1% (p = 0.794) |
| Hematological<br>conditions | 300 (1.6%) | 1.9% | 1.7% | 0.2% (p = 0.668) |
| Chronic kidney<br>disease | 232 (1.3%) | 1.9% | 1.7% | 0.2% (p = 0.725) |
| ***Uptake of preventive health services*** |  |  |  |  |
| HZ vaccination | 176 (1.0%) | 1.1% | 1.1% | 0.0% (p=0.991) |
| Statin use | 1,332 (7.2%) | 7.9% | 7.1% | 0.8% (p = 0.409) |
| Antihypertensive use | 1,089 (5.9%) | 7.1% | 5.6% | 1.4% (p = 0.087) |
| PPV | 3,443 (18.7%) | 21.1% | 19.4% | 1.7% (p = 0.216) |
| Influenza vaccination | 4,675 (25.4%) | 27.5% | 27.2% | 0.2% (p = 0.888) |
| DPT vaccination | 1,098 (6.0%) | 4.4% | 5.3% | -0.9% (p = 0.209) |
| Cancer screening | 491 (2.7%) | 1.0% | 1.5% | -0.6% (p = 0.150) |

### A one-week difference in age led to a large difference in HZ vaccination receipt

We estimated that adults born one week after the November 2 1936 date-of-birth eligibility cutoff had a 16.4 percentage point (95% CI: 13.2 – 19.5, p<0.001) higher probability of ever receiving the HZ vaccine than those born just one week earlier (**Figure S1**). Measured in the MSE-optimal bandwidth of 255 weeks around the November 2 1936 threshold, the mean HZ vaccination probability was 6.5% (95% CI: 5.6 – 7.3) versus 30.2% (95% CI: 29.0 – 31.4) among those ineligible versus those eligible for the vaccine, respectively.

In contrast to HZ vaccination uptake, we found no discontinuities across the November 2 1936 date-of-birth eligibility threshold in any of the following measures as assessed prior to the start date of the HZ vaccination program on November 1 2016: i) the probability of having received a diagnosis of any of the 15 most common diagnoses in our data; ii) uptake of preventive health services other than HZ vaccination; iii) diagnoses of dementia; and iv) risk factors for dementia on which we had information in our data (obesity, hyperlipidemia, hypertension, diabetes, current smoking, use of antihypertensive medications, and use of statins) (**Figure S1, S2** and **S12**). These tests, therefore, support the expectation that individuals born just on either side of the November 2 1936 date-of-birth threshold were exchangeable with each other except for a large difference in HZ vaccination uptake.

### The effect of eligibility for HZ vaccination on new diagnoses of dementia

Using our RD approach, we found that eligibility for free HZ vaccination decreased the probability of receiving a new dementia diagnosis over our 7.4-year follow-up period by 1.8 percentage points (95% CI: 0.4 – 3.3, p=0.013; **Figure 2**). The effect was similar across follow-up periods ranging from four to seven years, and grace periods ranging from zero to 156 weeks (**Figure 3**). There was no evidence of a significant treatment effect heterogeneity by gender (**Figure S3, Figure S4,** and **Text S3**).

**Fig. 2:**
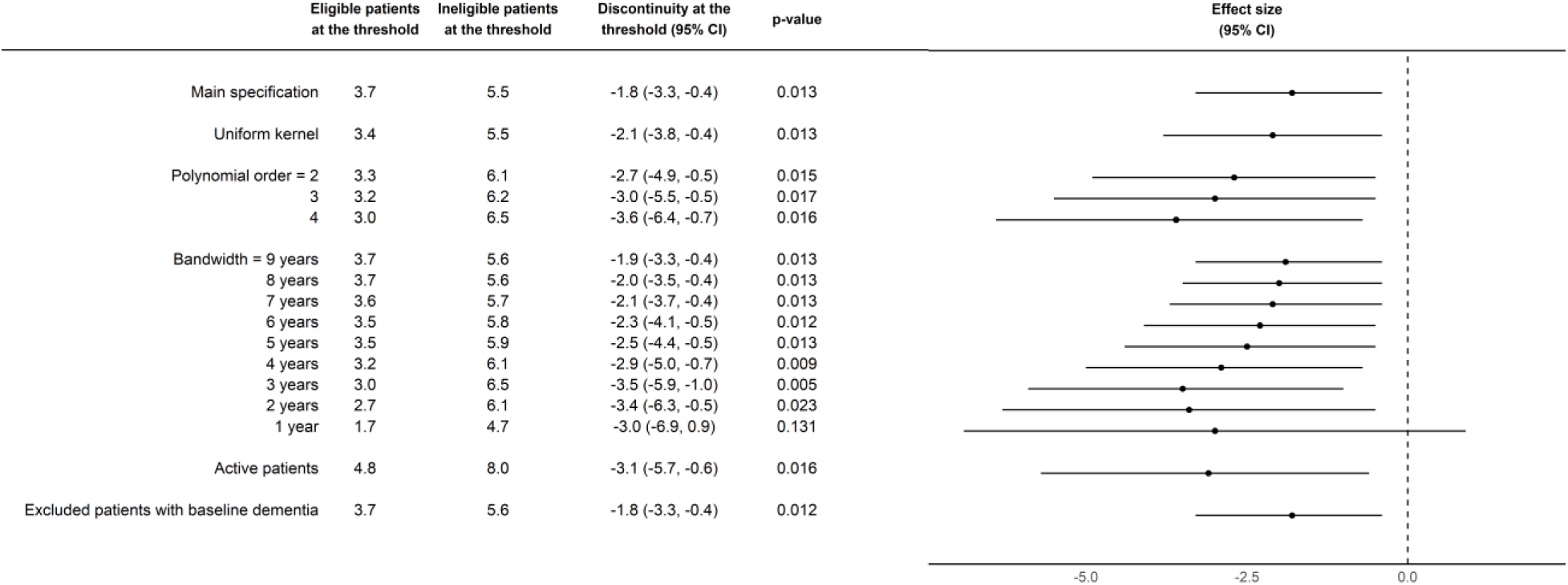
**The effect of being eligible for HZ vaccination on new diagnoses of dementia.^1,2^** Abbreviations: HZ = Herpes Zoster; CI = Confidence Interval; MSE = Mean Squared Error ^1^ Dots show the point estimate and horizontal bars the 95% confidence interval. ^2^ The main specification used triangular kernels, a local linear polynomial, and observations within the MSE-optimal bandwidth.

**Fig. 3:**
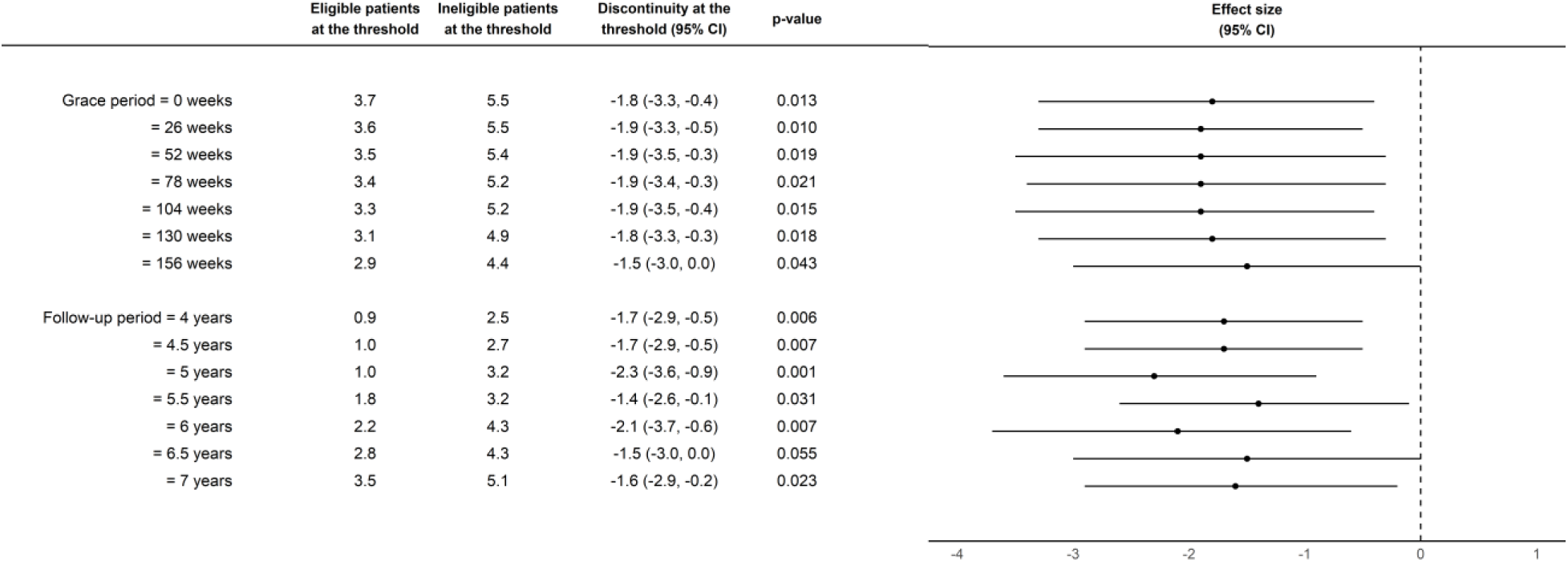
**The effect of being eligible for free HZ vaccination on new diagnoses of dementia, across different grace and follow-up periods.^1,2^** Abbreviations: HZ = Herpes Zoster; CI = Confidence Interval ^1^ With “grace periods” we refer to time periods since November 1 2016 after which follow-up time is considered to begin to allow for the time needed for a full immune response to develop after vaccine administration. ^2^ Horizontal bars depict 95% confidence intervals.

### Robustness checks

Our results were robust to a series of additional checks (**Figure 2**). First, the effect estimates remained similar in magnitude when using i) uniform kernel weights instead of triangular kernel weights, ii) local quadratic instead of local linear regression, and iii) bandwidths between one and nine years. We conducted similar robustness checks (shown in **Figures S5** and **S6**) for the effect of HZ vaccination eligibility on HZ vaccine uptake. Second, we also found a significant reduction in new diagnoses of dementia from HZ vaccination eligibility (−3.1 [95% CI: −5.7 – −0.6] percentage points, p=0.016]) when restricting our study cohort to the 61,903 frequent primary care visitors (“active” patients) in our data. Third, our results remained similar when excluding patients with a diagnosis of dementia recorded prior to the start date of the HZ vaccination program.

Fourth, we found that the acceleration factor was significantly larger than 1 (i.e., a protective effect from HZ vaccination eligibility) for all but the very shortest bandwidths (for which the 95% CIs are wide due to the smaller sample size) (**Figure S7**). Similarly, our Kaplan-Meier plots within a 12-, 9-, and 6-month bandwidth around the threshold, where eligible and ineligible patients are balanced on covariates (**Figure S10**), each showed that eligible patients take longer to be diagnosed with dementia than ineligible patients (**Figure S8**). Cumulative incidence curves with accompanying Gray’s tests for the same bandwidths (12, 9, and 6 months) also confirmed these findings (p = 0.01, p=0.01, and p=0.005, respectively) (**Figure S9**).

Finally, confirming the findings from our primary approach, our CRD approach found that HZ vaccination eligibility reduced the probability of a new diagnosis of dementia by 1.5 percentage points (95% CI: 0.2 – 2.7, p=0.020) over our 7.4-year follow-up period (**Figure S11**). These results were similar across different kernels, polynomial orders, bandwidths, and whether or not the study population was restricted to “active” patients and those without a dementia diagnosis at baseline.

### Testing for confounding

For our effect estimates to be unbiased, the key assumption that needs to be fulfilled is that no confounding variable changed abruptly at precisely the November 2 1936 date-of-birth eligibility threshold.^32,33^ Such a discontinuity of a confounding variable at the November 2 1936 date-of-birth threshold could occur if another intervention or policy used the identical date-of-birth threshold as its eligibility criterion as the HZ vaccination program. We investigated this possibility in three ways.

First, because another intervention that used a November 2 1936 date-of-birth eligibility criterion and was not specific to dementia would be unlikely to only affect dementia diagnoses without also having an impact on other common diagnoses, we investigated whether being eligible for HZ vaccination based on one’s date of birth had an effect on common disease diagnoses other than dementia. Using the same RD approach as in our primary analysis for dementia, we conducted this test for new diagnoses of each of the 15 most common diagnoses in the PenCS data. Unlike with dementia, being eligible for HZ vaccination based on one’s date of birth had no significant effect on the incidence of any of these 15 conditions over our 7.4-year follow-up period (**Figure 4**).

**Fig. 4:**
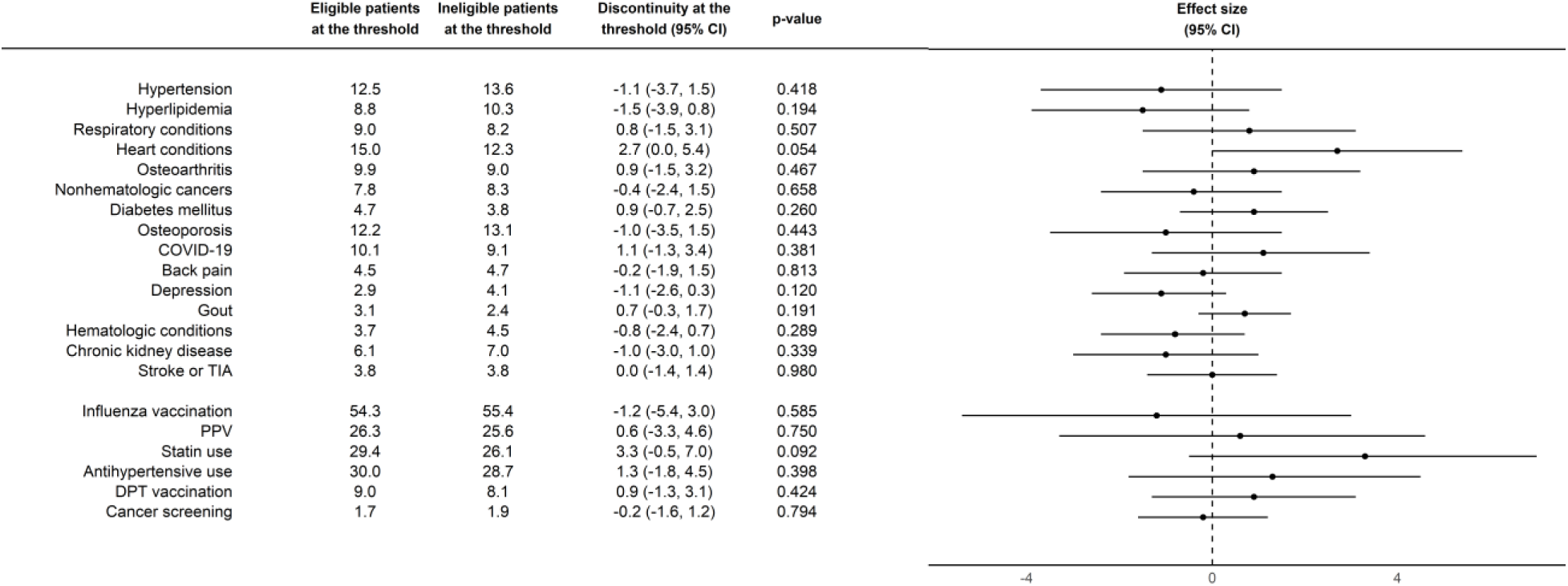
**Effect of being eligible for HZ vaccination on the 15 most common subsequent clinical diagnoses and uptake of other preventive health services during our 7.4-year follow-up period.^1,2,3^** Abbreviations: PPV = Pneumococcal Polysaccharide Vaccine; DPT = Diphtheria, Tetanus, and Pertussis; CI = Confidence Interval ^1^ Horizontal bars depict 95% confidence intervals. ^2^ The codes used to define each condition are shown in Table S1. ^3^ Cancer screening refers to the uptake of colorectal or breast cancer screening. As per Australian cancer screening guidelines, this was defined as uptake of fecal occult blood testing (for colorectal cancer screening) and mammography (for breast cancer screening).^56,57^

Second, we conducted the same analysis as for common clinical diagnoses shown in Figure 4 for indicators of preventive health services uptake. The rationale for these analyses was twofold: to investigate whether i) another intervention aimed at improving preventive health service use (e.g., another vaccination program) used a November 2 1936 date-of-birth eligibility criterion, and ii) HZ vaccination itself may have led to increased uptake of other preventive health services. For each of our indicators (influenza vaccination, pneumococcal vaccination, DPT vaccination, statin use, use of antihypertensive medications, and cancer screenings), we found no evidence that HZ vaccination eligibility affected preventive health services uptake (**Figure 4**).

Third, if another intervention used a November 2 date-of-birth eligibility criterion, then we may expect to see differences in the effect of this threshold on new diagnoses of dementia for birth years other than 1936. We, thus, implemented the same analysis as for our primary analysis (shown in Figure 2), but shifted the date-of-birth eligibility threshold to each of the three years before and after 1936. We found that the only date-of-birth threshold that resulted in a reduction in new diagnoses of dementia was the threshold used by the HZ vaccination program (i.e., November 2 1936; **Figure S13**).

## Discussion

This study found that individuals born immediately on either side of the November 2 1936 date-of-birth eligibility threshold for HZ vaccination had a large difference in their probability of receiving HZ vaccination whereas there was no difference between these individuals in past chronic disease diagnoses, preventive health services uptake, or dementia risk factors. Over the subsequent 7.4 years, being born immediately after November 2 1936 (and, thus, being eligible for HZ vaccination) versus being born immediately prior to November 2 1936 (and, thus, being ineligible) led to a large reduction in the probability of being diagnosed with dementia. We observed this effect only for dementia but not any other common diagnoses in our data. Our results were robust across a wide range of analytical specifications, as well as when using time-to-event models and restricting the study population to frequent primary care visitors.

In conjunction with our findings from a similar natural experiment in Wales,^9^ the results of our study suggest that HZ vaccination is a low-cost, high-reward intervention to reduce the burden of dementia. We believe that our findings call for investments into further research in this area, including clinical trials, further replications in other settings, populations, and health systems, and mechanistic research. Regarding mechanistic studies, several potential mechanisms have already been recognized. For example, reactivations of the varicella zoster virus have been linked to long-lasting cognitive impairment through vasculopathy,^37,38^ amyloid deposition and aggregation of tau proteins,^39^ neuroinflammation,^40–43^ as well as cerebrovascular disease resembling that seen in Alzheimer’s disease, including small to large vessel disease, ischemia, infarction, and hemorrhage.^40–45^ Additionally, there is a substantial body of evidence suggesting that the herpes simplex virus may contribute to the development of dementia,^2,46^ along with suggestive evidence that reactivations of the varicella zoster virus may lead to reactivations of the herpes simplex virus in the brain.^47^ Lastly, it is conceivable that HZ vaccination affects the dementia disease process through a pathogen-independent immunomodulatory pathway; a hypothesis that has been elaborated recently elsewhere.^48^

The key strength of this study is its quasi-experimental approach. Australia implemented its HZ vaccination program using a specific (maximum) date-of-birth eligibility threshold,^28^ which created population groups that differed in their age by only a minute degree but had large differences in the probability of receiving the HZ vaccine. The rollout of the HZ vaccine, therefore, created two comparison groups born just on either side of the eligibility who are likely to be similar to each other on observed and unobserved characteristics except for this difference in their probability of receiving HZ vaccination. Given our approach, a potentially confounding variable can only bias our findings if it changes abruptly at precisely the date-of-birth eligibility threshold that was used for the HZ vaccination program.^32,33^ Our tests found no evidence for the presence of such bias. It is important to note that our conclusions are also unlikely to be affected by ascertainment bias. If attending a primary care provider for HZ vaccination provided an opportunity for the health system to identify previously undetected cases of dementia, our analysis would underestimate, rather than overestimate, the vaccine’s effectiveness in reducing the incidence of new diagnoses of dementia. Additionally, if healthcare visits for herpes zoster episodes were an important way for the health system to identify previously undiagnosed chronic conditions, we would have expected to see effects of HZ vaccination eligibility on a wider range of common diagnoses beyond just dementia. We would have also expected a substantially smaller or absent effect of HZ vaccination on the incidence of dementia diagnoses among patients who frequently visit their primary care provider because one additional healthcare visit is presumably less likely to have an important influence on diagnosing previously undetected dementia in this population. We, however, found no such pattern.

The estimated effect size in our analysis was large in relative terms. However, it is important to recognize two limitations of our data when interpreting this effect size. First, the 95% confidence intervals around our estimates were comparatively wide, meaning that our data were compatible with considerably smaller effect sizes than our point estimates. The width of our confidence intervals may well also be the reason for which we did not observe the same gender effect heterogeneity as we have observed in our study in Wales.^9^ Second, there likely was substantial underdiagnosis of dementia in our data. For instance, an estimated 8.4% of all Australians over the age of 65 years are living with dementia,^49^ whereas only about 1.4% of patients in the PenCS data in the same age group in 2023 have been diagnosed with dementia. The underdiagnosis of conditions is a well-recognized limitation of working with primary care records from Australia, and not unique to dementia nor the PenCS data.^50–52^ Importantly, the degree of underascertainment of dementia is unlikely to differ between birth cohorts born just before versus just after November 2 1936.

Underreporting in our data was also the reason for which we refrained from scaling our effect estimates to the proportion of eligible patients who took up the vaccine. This would have allowed us to estimate the effect of actually receiving (as opposed to merely being eligible for) HZ vaccination. We reasoned that HZ vaccination is likely substantially underreported in our data because uptake of preventive health services in general appeared to be severely underreported. For instance, pneumococcal vaccination coverage (within the last five years) and influenza vaccination coverage (in the last year) among adults aged 65 years and older in Australia is thought to be approximately 55% and 75%, respectively.^53^ In our data, however, the corresponding percentages in this age group were only 21% and 33%, respectively. If the degree of underreporting of vaccinations was similar or larger for HZ vaccination as for influenza and pneumococcal vaccination, then any attempt to estimate the effect of receiving (as opposed to merely being eligible for) HZ vaccination using RD will greatly overestimate the effect of HZ vaccination receipt on dementia incidence. We, therefore, chose to only analyze the effect of being eligible for HZ vaccination.

Our study has several additional limitations. First, our analysis only provided “local” estimates of the effect of HZ vaccination on the incidence of dementia, i.e., estimates for patients who were approximately 79 and 80 years old at the time of the start of the HZ vaccination program.

Second, given that we had data from a non-random sample of primary care practices in Australia, our dataset was unlikely to be representative of all primary care patients in the country. Third, because the recombinant subunit HZ vaccine (Shingrix [GSK]) was covered by the NIP starting only on November 1 2023,^55^ our effect estimates apply to the live-attenuated HZ vaccine (Zostavax [Merck]) only. Fifth, our sample restrictions, including the removal of patients with missing IDs, might introduce a spurious association between eligibility for the HZ vaccine and dementia, especially if we are conditioning on colliders, or variables that are influenced both by our treatment and outcome variables. However, these collider variables would have to change abruptly at the November 2 1936 date-of-birth threshold to introduce bias into our analysis. Lastly, the MSE-optimal bandwidth – 482 weeks – that we adopted for our RD analysis on dementia is relatively large. However, within this bandwidth, our analysis assigned higher weights to individuals born most closely around the November 2 1936 eligibility threshold. In addition, our results were not substantially different when we adopted narrower bandwidths. Our estimates remained negative and statistically significant for bandwidths as small as 2 years around the threshold, and they remained statistically significant using 90% confidence intervals with a 1.5 year bandwidth. Even with a 1-year bandwidth, though we lost significance because of a substantially smaller sample size, our point estimates were nearly unchanged. Most importantly, regardless of the size of the bandwidth, our RD design merely assumed that among patients in our bandwidth there do not exist confounding variables that change abruptly at precisely the November 2 1936 date-of-birth threshold.^32,33^

In conclusion, corroborating our findings from a similar natural experiment in Wales,^9^ we found that being eligible for HZ vaccination based on one’s date of birth significantly reduced the incidence of new dementia diagnoses over a 7.4-year follow-up period. Due to their ability to compare individuals who had large differences in their probability of receiving HZ vaccination merely because of being born somewhat earlier or later, this study and our analysis in Wales provide evidence that is more robust to confounding concerns (e.g., healthy vaccinee bias) than that of the existing associational evidence.

## Data Availability

The data that support the findings of this study are available from PenCS. Researchers must request access to the data directly from PenCS. The authors have no permission to share the data. All codes to define variables are available in the Supplement. All statistical analysis code (in R) will be made available in a publicly accessible GitHub repository upon acceptance of the manuscript for publication.

## Acknowledgments

The authors are grateful for the continuous advice and support from the PenCS team who made this data available for research.

## Authors’ contributions

M.P. devised the methodology, analyzed and processed the data, created data visualizations, interpreted the results, wrote the original draft, and reviewed and edited the original draft. C.B. devised the methodology, interpreted the results, and reviewed and edited the original draft. F.P. and F.M. interpreted the results, and reviewed and edited the original draft. R.P. interpreted the results, and reviewed and edited the original draft. P.G. conceived the overall project, acquired funding, conceived the study, devised the methodology, was responsible for administration and supervision, interpreted the results, wrote the original draft, and reviewed and edited the original draft.

## Data Access, Responsibility, and Analysis

M.P. had full access to all the data in the study and takes responsibility for the integrity of the data and the accuracy of the data analysis.

## Data sharing

The data that support the findings of this study are available from PenCS.^33^ Researchers must request access to the data directly from PenCS. The authors have no permission to share the data. All codes to define variables are available in the Supplement. All statistical analysis code (in R) will be made available in a publicly accessible GitHub repository upon acceptance of the manuscript for publication.

