## Supplement for "Effect of herpes zoster vaccination on dementia occurrence: A quasi-experimental study in Australia"

**The file includes:**

Table S1
Text S1
Text S2
Text S3
Figures S1 to S14

**Table S1: Open-ended texts for diagnoses in PenCS data**

**Text S1: Data source**
**Text S2: Statistical analysis**
**Text S3: Regression equations**

**List of Figures**

Figure S1: The date-of-birth eligibility cutoff led to a large discontinuity in HZ vaccination receipt but there is no such jump at the cutoff in the uptake of other preventive interventions.

Figure S2: Baseline balance checks using the 15 most common diagnoses in the PenCS data and indicators of preventive health services uptake prior to the start of the vaccination program

Figure S3: Effect of being eligible for herpes zoster vaccination on new diagnoses of dementia over a 7.4-year follow-up period, by gender

Figure S4: The discontinuity in HZ vaccine uptake was similar between women (panel A) and men (panel B).

Figure S5: The effect of being eligible for herpes zoster vaccination on herpes zoster vaccination uptake using different kernel weights and functional forms

Figure S6: The effect of being eligible for herpes zoster vaccination on herpes zoster vaccination uptake using different bandwidths

Figure S7: The acceleration factor across different bandwidths, which indicates how much longer patients eligible for HZ vaccination take to be diagnosed with dementia than ineligible patients

Figure S8: Survival functions (Kaplan-Meier plots) for patients eligible and ineligible for HZ vaccination

Figure S9: Cumulative incidence curves for patients eligible and ineligible for HZ vaccination

Figure S10: Differences in covariates between patients eligible and ineligible for HZ vaccination, for one-year, nine-month, and six-month bandwidths

Figure S11: Effect of being eligible for herpes zoster vaccination on new diagnoses of dementia over a 7.4-year follow-up period, using the comparative regression discontinuity design

Figure S12: There was balance at baseline across the November 2 1936 date-of-birth threshold in dementia risk factors and dementia diagnoses recorded prior to the start of the herpes zoster vaccination program

Figure S13: The November 2 date-of-birth threshold only had a protective effect for the occurrence of dementia in the birth year (1936) that was used by the HZ vaccination program as eligibility criterion

Figure S14**:** There was balance at baseline across the November 2 1936 date-of-birth threshold in active patient status and missing dementia diagnosis dates, but not for missing patient IDs

| Variables | Text Codes |
|---|---|
| Dementia | Alzheimer's Disease; Dementia; Dementia, Early Onset; Dementia, Vascular; Frontotemporal Dementia; Lewy Body Dementia; Vascular Dementia |
| Herpes zoster (HZ) vaccine | Zostavax; Shingrix |
| Influenza vaccine | Aflqua; Afluria Quad; Afluria Quad (NIP); Afluria Quad (Non-NIP); Afluria Quad (Non NIP); Afluria Quad NIP; Agrippal; Arxflu; Biocsl Fluvax; Fluad; Fluad-Quad; Fluad Quad; Fluarix; Fluarix Tetra; Fluarix Tetra (NIP); Fluarix Tetra (Non-NIP); Fluarix Tetra (Non NIP); Flucelvax Quad; Fluquadri; Flurivin; Fluvax; Fluzone High-Dose; Fluzone High-Dose Quad; Generic Influenza; H1N1 Influenza; Influenza; Influenza (Quadrivalent); Influvac; Influvac 2011; Influvac Tetra; Intanza; Intanza 9mcg; Panvax; Panvax H1N1; Quadaf; Vaxigrip; Vaxigrip Tetra |
| Pneumococcal Polysaccharide Vaccine (PPV) | Pneumococcus (13 Valent); Prevenar; Prevenar 13; Pneumococcal; Pneumococcus (23 Valent); Pneumovax; Pneumovax 23 |
| Statins | Amlodipine (As Besilate)/Atorvastatin (As Calcium); Amlodipine Besylate, Atorvastatin; Amlodipine, Atorvastatin; Amlodipine/Atorvastatin; Atorvastatin; Atorvastatin (As Calcium Trihydrate); Atorvastatin (As Calcium); Ezetimibe, Atorvastatin; Ezetimibe, Rosuvastatin; Ezetimibe, Simvastatin; Ezetimibe/Atorvastatin (As Calcium Trihydrate); Ezetimibe/Rosuvastatin; Ezetimibe/Rosuvastatin (As Calcium); Ezetimibe/Simvastatin; Fluvastatin; Pravastatin Sodium; Rosuvastatin; Rosuvastatin (As Calcium); Rosuvastatin 10mg Coated Tablet; Rosuvastatin 20mg Coated Tablet; Simvastatin |
| Diphtheria-Tetanus-Pertussis vaccine | Acellular DTP; Adacel; Adacel Polio; Boostrix; Boostrix-IPV; Boostrix IPV; DTPA; Infanrix; Infanrix IPV; Quadracel; Tripacel |
| Antihypertensive medications | Amiloride Hydrochloride/Hydrochlorothiazide; Amlodipine; Amlodipine (As Besilate); Amlodipine (As Besilate)/Atorvastatin (As Calcium); Amlodipine (As Besilate)/Valsartan; Amlodipine Maleate; Atenolol; Bisoprolol Fumarate; Candesartan Cilexetil, Hydrochlorothiazide; Carvedilol; Chlortalidone; Diltiazem Hydrochloride; Enalapril Maleate, Hydrochlorothiazide; Felodipine; Hydralazine Hydrochloride; Hydrochlorothiazide; Indapamide Hemihydrate; Irbesartan, Hydrochlorothiazide; Labetalol Hydrochloride; Lercanidipine Hydrochloride; Lercanidipine Hydrochloride/Enalapril Maleate; Metoprolol Succinate; Metoprolol Tartrate; Minoxidil; Nebivolol (As Hydrochloride); Nifedipine; Olmesartan Medoxomil, Hydrochlorothiazide; Perindopril Arginine, Indapamide Hemihydrate; Propranolol Hydrochloride; Ramipril, Felodipine; Spironolactone; Telmisartan, Hydrochlorothiazide; Trandolapril, Valsartan/Hydrochlorothiazide; Verapamil; Verapamil Hydrochloride |
| Back pain | Back Ache; Back Muscle Strain; Back Pain; Back Pain - Lumbar; Back Pain - Lumbo-Sacral; Back Pain - Sacral; Backache; Chronic Back Pain; Chronic Low Back Pain; Low Back Pain; Low Back Pain - Mechanical; Lumbago; Lumbar - Pain; Lumbar Back Injury; Lumbar Back Muscle Strain; Lumbar Sprain; Mechanical Back Pain; Sacral Spinal Pain |
| Osteoarthritis | Ankle Osteoarthritis; Arthritis - Osteo; Bilateral Osteoarthritis Of Knee; Cervical - Osteo Arthritis; Cervical Spine Osteoarthritis; Hip Osteoarthritis; Hip Osteoarthrosis; Knee Osteoarthritis; Lumbar - Osteo Arthritis; Lumbar Spine Osteoarthritis; Osteoarthritis; Osteoarthritis - Fingers; Osteoarthritis - Glenohumeral Joint; Osteoarthritis - Hands; Osteoarthritis - Neck; Osteoarthritis - Shoulder; Osteoarthritis - Spine; Osteoarthritis Of Hand; Spondylosis |
| Hypertension | Essential Hypertension; High Blood Pressure; HT (Hypertension); Hypertension - Controlled; Hypertension - Isolated Systolic; Hypertension - Labile; Hypertension, Essential; Labile Blood Pressure |
| Osteoporosis | Osteopenia; Osteoporosis; Osteoporosis – Corticosteroid Induced; Osteoporosis – No Fracture; Osteoporosis With Fracture; Osteoporosis, Steroid Induced |
| Hyperlipidemia | Dyslipidemia; Familial Hypercholesterolemia; High Cholesterol; Hypercholesterolemia; Hyperlipidemia; Hyperlipidemia – Controlled; Hyperlipidemia Type 2; Hypertriglyceridemia |

| | |
|---|---|
| Non-hematological cancers | Adenocarcinoma – Colon; Adenocarcinoma – Prostate; Adenocarcinoma Of Lung; Adenocarcinoma Of Prostate; Anal Carcinoma; Basal Cell Carcinoma; Basal Cell Carcinoma – Nodular; Basal Cell Carcinoma, Superficial; BCC – Infiltrative; Bladder Cancer; Bone Metastases; Bowel Cancer; Bowen's Disease; Brain Metastases; Brain Tumor; Breast Cancer; Breast Cancer, Lobular; Breast Carcinoma; Caecal Carcinoma; Cancer; Carcinoid Tumor; Carcinoma – Prostate; Carcinoma Of Skin; Carcinoma Of The Bladder; Carcinoma Of The Breast; Carcinoma Of The Colon; Carcinoma Of The Prostate; Cervical Cancer; Cervical Carcinoma; Cholangiocarcinoma; Colon – Adenocarcinoma; Colon Carcinoma; Colonic Cancer; Colonic Carcinoma; Ductal Carcinoma In Situ; Ductal Carcinoma Infiltrating; Dysplastic Naevus Syndrome; Endometrial Adenocarcinoma; Gastric Cancer; GIST; Hepatocellular Carcinoma; Intraepidermal Carcinoma; Intraepithelial Carcinoma Of Skin; Kerato-Acanthoma; Keratoacanthoma; Kidney Cancer; Laryngeal Carcinoma; Lentigo Maligna Melanoma; Liver Cancer; Liver Metastases; Lobular Breast Cancer; Lung Cancer; Lynch Syndrome; Malignant Melanoma; Melanoma; Melanoma – Invasive; Melanoma In Situ; Melanoma, Nodular; Melanoma, Superficial Spreading; Meningioma; Meningioma, Cranial; Mesothelioma; Metastases, Bone; Metastatic Cancer; Metastatic Carcinoma; Metastatic Melanoma; Nasopharyngeal Carcinoma; Nodular BCC; Oesophageal Cancer; Oesophageal Carcinoma; Ovarian Cancer; Ovarian Carcinoma; Pancreatic Cancer; Pancreatic Tumor; Papillary Thyroid Carcinoma; Parotid Carcinoma; Phaeochromocytoma; Prostate Cancer; Prostate Carcinoma; Prostatic Adenocarcinoma; Rectal Adenocarcinoma; Rectal Cancer; Rectal Carcinoma; Renal Adenocarcinoma; Renal Cancer; Renal Carcinoma; Sarcoma; SCC (Squamous Cell Carcinoma); Skin Cancer; Skin Cancers – Multiple; Small Cell Carcinoma Of Lung; Squamous Cell Carcinoma; Squamous Cell Carcinoma In-Situ; Squamous Cell Carcinoma Of Skin; Superficial Multifocal BCC; Superficial Spreading Melanoma; Testicular Cancer; Thyroid Cancer; Thyroid Carcinoma; Tongue Carcinoma; Tonsillar Carcinoma; Transitional Cell Carcinoma – Bladder; Transitional Cell Carcinoma Of The Bladder; Urothelial Cell Carcinoma Of The Bladder; Uterine Adenocarcinoma |
| COVID-19 infection | Covid-19; Covid-19 (Coronavirus) Infection; Covid-19 (Coronavirus) Infection Mild |
| Depression | Depressed; Depressed Mood; Depression; Depression – Endogenous; Depression – Major; Depression – Reactive; Depression With Melancholic Features; Depression, Melancholic; Depression, Non Melancholic; Depressive Episode, Major; Dysthymia; Major Depressive Episode; Premenstrual Dysphoric Disorder |
| Gout | Gout; Gout – Acute Attack; Gouty Arthritis; Gouty Tophi; Gouty Tophus; Hyperuricemia; Tophaceous Gout |
| Stroke or Transient Ischemic Attack (TIA) | Cerebellar Infarct; Cerebral Haemorrhage; Cerebral Infarction; Cerebrovascular Accident; CVA (Cerebrovascular Accident); Haemorrhagic CVA; Haemorrhagic Stroke; Intracerebral Haemorrhage; Intracranial Haemorrhage; Ischemic Stroke; Lacunar Infarct; Lacunar Stroke; Stroke; Stroke - Ischemic; Stroke, Haemorrhagic; Stroke, Ischemic; Subarachnoid Haemorrhage; TIA (Transient Ischemic Attack) |
| Hematological conditions | AML; Anemia; Anemia – Blood Loss; Anemia – Iron Deficiency; Anemia Due To Blood Loss; Anemia Of Chronic Disease; Anemia, Microcytic; Anemia, Normocytic Normochromic; B12 Deficiency Anemia; Beta Thalassemia Trait; Chronic Lymphatic Leukemia; Chronic Lymphocytic Leukemia; Chronic Myeloid Leukemia; CLL; Essential Thrombocytosis; Hodgkin's Lymphoma; Leukemia; Low Hemoglobin; Lymphoma; Macrocytic Anemia; Mantle Cell Lymphoma; Monoclonal B Cell Lymphocytosis; Multiple Myeloma; Myelo-Dysplastic Syndrome; Myelodysplasia; Myeloproliferative Disease; Normocytic Normochromic Anemia; Pernicious Anemia; Polycythaemia Rubra Vera; Thalassemia; Thalassemia Minor; Thalassemia Trait; Thrombocythaemia; Waldenstrom's Macroglobulinaemia |
| Heart disease | Acute Coronary Syndrome; Acute Myocardial Infarction; AF; AF (Atrial Fibrillation); AMI; AMI (Acute Myocardial Infarction); Angina; Angina Pectoris; Angina, Stable; Angina, Unstable; Aortic Incompetence; Aortic Regurgitation; |

| | |
|---|---|
| | Aortic Root Dilatation; Aortic Sclerosis; Aortic Stenosis; Aortic Stenosis, Valvular; Aortic Valve Disease, Mixed; Aortic Valve Stenosis; Arrhythmia; Atrial Ectopic Beats; Atrial Fibrillation; Atrial Fibrillation - Paroxysmal; Atrial Fibrillation, Non-Valvular; Atrial Fibrillation, Valvular; Atrial Flutter; Atrial Septal Defect; Atrial Tachycardia; AV Block; Bicuspid Aortic Valve; Bifascicular Block; Bigeminy; Bradyarrhythmia; Bradycardia; Cardiac Arrhythmia; Cardiac Failure; Cardiomegaly; Cardiomyopathy; Cardiomyopathy - Dilatation; Cardiomyopathy, Dilated; Cardiomyopathy, Ischemic; Cardiovascular Disease; CCF; Complete Heart Block; Congestive Cardiac Failure; Congestive Heart Failure; Cor Pulmonale; Coronary Artery Calcification; Coronary Artery Disease; Coronary Artery Spasm; Coronary Heart Disease; Diastolic Cardiac Dysfunction; Diastolic Dysfunction; Diastolic Heart Failure; Dilated Aortic Root; Dilated Cardiomyopathy; Ectopics, Atrial; First Degree Heart Block; Fistula, Arteriovenous; Heart Attack; Heart Block; Heart Block, First Degree; Heart Block, Second Degree; Heart Disease; Heart Failure; Heart Failure - Preserved Ejection Fraction; Heart Failure - Reduced Ejection Fraction; HFPEF; HFREF; Hypertrophic Cardiomyopathy; IHD; IHD (Ischemic Heart Disease); Infarction, Myocardial; Infective Endocarditis; Inferior Myocardial Infarction; Ischemic Cardiomyopathy; Ischemic Heart Disease; Left Bundle Branch Block; Left Ventricular Diastolic Dysfunction; Left Ventricular Dysfunction; Left Ventricular Failure; Left Ventricular Hypertrophy; Left Ventricular Systolic Dysfunction; LVF; LVF (Left Ventricular Failure); MI; Mitral Incompetence; Mitral Prolapse; Mitral Regurgitation; Mitral Stenosis; Mitral Valve Disease, Mixed; Mitral Valve Prolapse; Myocardial Infarction; Myocardial Infarction - Inferior; Myocardial Infarction, Non STEMI; Myocardial Infarction, STEMI; Myocarditis; Non-St-Elevation Myocardial Infarction (NSTEMI); Non St Elevation Myocardial Infarction; NSTEMI; NSTEMI (Non-St-Elevation Myocardial Infarction); Paroxysmal Atrial Fibrillation; Paroxysmal Atrial Tachycardia; Pericardial Effusion; Pericarditis; Premature Ventricular Contractions; Rapid Atrial Fibrillation; RBBB; Rheumatic Fever; Rheumatic Heart Disease; Right Bundle Branch Block; Sick Sinus Syndrome; Sinus Arrhythmia; Sinus Bradycardia; Sinus Tachycardia; Stable Angina; STEMI; STEMI (St-Elevation Myocardial Infarction); Supraventricular Tachycardia; SVT; Syncope, Cardiac; Tachycardia; Takotsubo Cardiomyopathy; Tricuspid Incompetence; Tricuspid Regurgitation; Unstable Angina; Valvular Disease Of Heart; Valvular Heart Disease; Ventricular Bigeminy; Ventricular Ectopics; Ventricular Extrasystoles; Ventricular Tachycardia; VT |
| Chronic kidney disease | Chronic Kidney Disease; Chronic Kidney Disease – Stage 2; Chronic Kidney Disease – Stage 3; Chronic Kidney Disease – Stage 4; Chronic Kidney Disease, Stage 1; Chronic Kidney Disease, Stage 2; Chronic Kidney Disease, Stage 3a; Chronic Kidney Disease, Stage 3b; Chronic Kidney Disease, Stage 5; Chronic Renal Failure; Diabetic Nephropathy; Kidney Failure, Chronic; Renal Failure, Chronic; Renal Impairment |
| Diabetes mellitus | Diabetes; Diabetes – Controlled; Diabetes – Unstable; Diabetes Mellitus; Peripheral Vascular Disease, Diabetic; Diabetic Foot; Diabetic Foot Ulcer; Diabetic Ketoacidosis; Diabetic Nephropathy; Diabetic Neuropathy; Diabetic Retinopathy; Diabetic Retinopathy – Non Proliferative; Foot Ulcer, Diabetic; Gastroparesis, Diabetic; Diabetes Mellitus – Type I; IDDM (Insulin Dependent Diabetes Mellitus); Diabetes Mellitus – NIDDM; Diabetes Mellitus – Type II; Diabetes Mellitus, Type 2; Diabetes Type II Requiring Insulin; NIDDM; NIDDM (Non Insulin Dependent Diabetes Mellitus); Type 2 Diabetes; Type 2 Diabetes Mellitus |
| Respiratory conditions | Acute Exacerbation Of COPD; Allergic Asthma; Asbestosis; Aspergillus Pneumonia; Aspiration Pneumonia; Asthma; Asthma - Allergy Induced; Asthma, Childhood; Asthma - Chronic Persistent; Asthma - Exercise Induced; Asthma, Frequent Episodic; Asthma - Infrequent Episodic; Asthma Review; Asthma, Allergic; Asthma, Allergy Induced; Asthma, Infective Exacerbation; Atelectasis; Bronchial Asthma; Bronchiectasis; Bronchitis, Bacterial, Protracted; Chronic Bronchitis; Chronic Obstructive Airways Disease; Chronic Obstructive Pulmonary Disease; COAD - Infective Exacerbation; COAD (Chronic Obstructive Airways Disease); COAD, Infective Exacerbation; |

| | |
|---|---|
| | COPD; COPD - Infective Exacerbation; COPD (Chronic Obstructive Pulmonary Disease); COPD, Infective Exacerbation; Emphysema; Empyema; Fibrosis Of Lung; Idiopathic Pulmonary Fibrosis; Infective Exacerbation Of Asthma; Infective Exacerbation Of COAD; Infective Exacerbation Of COPD; Infective Exacerbation Of Chronic Bronchitis; Interstitial Lung Disease; Interstitial Pulmonary Fibrosis; Lobectomy; Lung Abscess; Lung Collapse; Lung Lobectomy; Obstructive Sleep Apnea; Pleural Effusion; Pneumonectomy; Pneumothorax; Pneumothorax – Spontaneous; Pneumothorax, Traumatic; Pulmonary Embolism; Pulmonary Fibrosis; Pulmonary Tuberculosis; Respiratory Failure; Restrictive Lung Disease; Sarcoidosis; Sleep Apnea; Smokers' Cough; TB (Tuberculosis); Tuberculosis Of The Lung; Wheezy Bronchitis |

**Table S1: Open-ended texts for diagnoses in PenCS data.**[1]

[1] This table provides the open-ended texts for each diagnosis analyzed in this paper.

**Text S1: Data source:**
We used data from PenCS,[1] an Australian-owned health informatics company, which provides detailed primary care electronic health records to researchers. The data included diagnoses, immunizations and other healthcare procedures, as well as prescribed medications from 65 general practitioner (GP) practices across each of Australia's six states and the Australian Capital Territory. These were GP practices that voluntarily consented to share their electronic health record data for research. 27 GP practices were located in New South Wales, 17 in Queensland, one in South Australia, one in Tasmania, 14 in Victoria, four in Western Australia, and one in the Australian Capital Territory. As classified by the Modified Monash Model,[2] 14 practices were in a metropolitan area, 40 in regional centers, 10 in small rural towns, and one in a remote community. The practices ranged in size from approximately 200 to 4,000 patients, with the average practice having 1,672 patients. Relative to practices in Australia more generally, our data under-samples practices in metropolitan areas (where the plurality of practices in Australia are located).[3] Moreover, the distribution of practices across states/territories does not exactly mirror the general population – for instance, the PenCS data has 14 practices in Victoria and 17 in Queensland, even though there are more GPs working in Victoria than Queensland.[4]

General practitioners function as gatekeepers in the Australian healthcare system such that patients generally only qualify for the Medicare Benefits Schedule for specialist care after a referral from a GP.[5] For the purposes of our analysis, PenCS provided us with patients' dates of birth in weeks. As is customary in Australia's primary care records, diagnoses were coded by PenCS using open-ended text fields provided by the GP. The text fields used to define each diagnosis in our analysis are listed in **Table S1**. PenCS does not link any of its primary care records to hospital records or mortality registers.

This study did not use the MedicineInsight database because it does not provide date of birth at a more granular level than years and is not currently available for research.[6,7]

One limitation of our dataset is that there was significant underreporting of clinical diagnoses and vaccination uptake. For instance, an estimated 8.4% of all Australians over the age of 65 are living with dementia,[8] whereas only about 1.4% of patients in the PenCS data in the same age group in 2023 have been diagnosed with dementia. The underdiagnosis of conditions is a well-recognized limitation of working with primary care records from Australia, and not unique to dementia nor the PenCS data.[9-11] Importantly, the degree of underascertainment of dementia is unlikely to differ between birth cohorts born just before versus just after November 2 1936.

We also reason that HZ vaccination was likely substantially underreported in our data because uptake of preventive health services in general appeared to be severely underreported. For instance, pneumococcal vaccination coverage (within the last five years) and influenza vaccination coverage (in the last year) among adults aged 65 years and older in Australia is thought to be approximately 55% and 75%, respectively.[12] In our data, however, the corresponding percentages in this age group were only 21% and 33%, respectively.

**Text S2: Statistical Analysis**

*Regression discontinuity:*

Our analysis was based on the rationale that individuals born very close to either side of the November 2 1936 threshold are expected to be similar to each other in observed and unobserved characteristics except for their eligibility status for HZ vaccination. We tested for differences in our outcomes at the November 2 1936 date-of-birth eligibility threshold for HZ vaccination using regression discontinuity (RD) analysis, which is a well-established statistical technique for causal effect estimation.[13] Regression discontinuity provides an unbiased effect estimate as long as any confounding variables do not abruptly change at exactly the November 2 1936 threshold.[14,15] This assumption was unlikely to be violated in this study because there was, to our knowledge, no other relevant policy or intervention that used the identical date of birth threshold as its eligibility criterion as the HZ vaccination program. As described below, we conducted a series of tests to further substantiate that this assumption was met.

As per recommended practice,[13-16] we used local linear regression with triangular kernel weights on observations within a mean squared error (MSE)-optimal bandwidth of the date-of-birth eligibility threshold. In robustness checks, we also implemented our analysis using different bandwidth choices, local quadratic regression, and uniform weights. Local linear regression is the recommended and most robust approach for RD analyses even in situations in which the relationship between the assignment variable (here, date of birth) and the outcome is exponential.[16] Triangular kernel weights give more weight to those observations closer to the eligibility threshold and less weight to observations further away.[15] The MSE-optimal criterion is used as an objective criterion to balance precision and bias in estimation.[15] We calculated the MSE-optimal bandwidth for each analysis separately.

We tested for an effect heterogeneity by gender by i) performing the analysis separately among women and men, and ii) running an interaction model that measured the difference in effects between men and women. The regression equations for all analyses are provided in **Text S1**.

Given its implementation using local linear regression, the effect estimates obtained in RD are absolute effect estimates. We, thus, consistently reported the absolute (in percentage points) rather than the relative effects of HZ vaccination eligibility.

*Survival analysis:*

In additional analyses, we also accounted for the different amounts of follow-up time across patients. Our first time-to-event approach was a cause-specific accelerated failure time (AFT) model, using the approach by Adeleke et al.,[17] who have specifically adapted this model to RD settings. Importantly, the AFT model assumes that the survivor functions for patients who are eligible for HZ vaccination and those who are ineligible can be characterized by $S(\varphi t \mid \text{eligible}) = S(t \mid \text{eligible})$, where $\varphi$ denotes the "acceleration factor." Note that the acceleration factor has the opposite interpretation from the hazard ratio; $\varphi > 1$ implies that the amount of time for eligible patients to experience the event (i.e., receive a dementia diagnosis) is longer than for ineligible patients. To measure follow-up time for each patient, we used the difference in time between November 1 2016 and the date of their dementia diagnosis (the date of the patient's most recent consultation with their GP) for patients diagnosed (undiagnosed) with dementia. For each of our survival analysis approaches, we omitted patients who either had dementia at baseline (i.e., had received a dementia diagnosis prior to November 1 2016) or who had completely missing consultation data.

Survival analysis methods, including our first approach described above, typically assume that any censoring in the data is uninformative. This is not always the case, however, especially in the presence of competing risks – when the patient experiences another event that alters the probability that a patient experiences the primary event of interest (i.e., being newly diagnosed with dementia). We therefore performed additional tests by appealing to the “local randomization” approach to the RD design. That is, within a narrow bandwidth around the threshold, individuals are likely similar to each other and, thus, the data within such a narrow bandwidth can be analyzed like in a randomized trial. More detail on this alternative approach to RD is provided elsewhere.[14,18] Using this notion of local randomization, we employed a second approach that enables us to use other, more traditional, time-to-event methods. First, we shortened the bandwidth to a narrow area around the date-of-birth eligibility threshold, such that it is more likely that patients below and above the threshold are similar in terms of observed and unobserved characteristics, even without controlling for the running variable (local randomization). Then, we used methods that are commonly used in analyses of time-to-event data: (1) we created Kaplan-Meier plots for each treatment condition; and (2) we compared cumulative incidence curves between eligible and ineligible patients using Gray’s test. This approach assumed that, within the narrow bandwidths used in our analyses, patients born before versus after the date-of-birth eligibility threshold are exchangeable with each other.

Many studies have documented an inverse relationship between cancer and dementia diagnoses.[19-21] Beyond the possible common overlap in the pathogenesis of dementia and cancer, there are a few postulated methodological biases that may contribute to this empirical finding, including competing risks of death – i.e., cancer patients, who have higher mortality risk, are less likely to be diagnosed with dementia. Moreover, survival biases may arise if unmeasured or unknown factors affect both cancer survival and dementia diagnoses. Finally, diagnostic biases may either bias in favor or against an inverse relationship. For instance, cancer diagnoses may cause GPs to overlook dementia symptoms and thus create delay in dementia diagnoses, but on the other hand, cancer patients may have more contact with their doctor, which could accelerate the timing of dementia diagnoses. We note, however, that these biases are unlikely to vary precisely at the date-of-birth eligibility threshold, such that these methodological issues are unlikely to be a concern for our identifying assumptions.

*Comparative regression discontinuity:*
One common drawback of RD analyses is the often relatively high level of imprecision in estimating effects at the threshold.[22] To improve precision, we used a variant of the RD design, called comparative RD (CRD), in secondary analyses, by leveraging an additional source of untreated data from older patients in our sample. By adding these data, CRD tends to provide increased statistical power relative to standard RD.[22] This improved statistical efficiency can yield point estimates that are more similar to those from randomized controlled trials than those from standard RD.[23,24]

In addition to the main cohort of patients who were born in close proximity to the date of birth-based eligibility threshold, our CRD design analyzed a comparison cohort of patients that was always ineligible for HZ vaccination. As a result, in our CRD, there were two sets of vaccine-ineligible individuals: i) ineligible patients in the main cohort (i.e., those born before the date of birth-based eligibility threshold); and ii) patients in the comparison cohort. For our comparison cohort, we used the youngest cohort of patients that was older (and thus always ineligible) than the patients born within the MSE-optimal bandwidth around the November 2 1936 eligibility threshold for HZ vaccination. We defined our comparison cohort using the same MSE-optimal bandwidth size as we used for ineligible patients in our main cohort. As such, this cohort was born (approximately) between May 13 1918 and August 1 1927, whereas the main cohort was born approximately between August 8 1927 and January 28 1946. To account for the age difference between the main and comparison cohort, we moved the follow-up period for identifying new dementia diagnoses for the comparison cohort earlier, so that dementia diagnoses for ineligible patients in both cohorts are measured within the same age timeframe. Specifically, the follow-up period for the comparison cohort was from August 1 2007 to January 1 2015. We implemented our CRD analysis using local linear regression and uniform kernel weights.

Our CRD approach assumed that the relationship between age and dementia incidence was similar for patients in our comparison cohort as for patients ineligible for HZ vaccination in our main cohort. To evaluate this assumption, we compared the trends of age with the incidence of new dementia diagnoses within the MSE-optimal bandwidth around the November 2 1936 eligibility threshold between patients in the comparison cohort and patients ineligible for HZ vaccination in the main cohort. In addition to visual inspection, we did this by testing whether the age-dementia trends were statistically significantly different from each other between these two patient groups using regression analysis. Details on these regressions are provided in **Text S3**. We found no evidence that the assumption for the valid use of CRD was violated (p = 0.387).

*Baseline balance checks:*
The robustness of our study design to confounding rests on the intuition that potential confounding variables are unlikely to change abruptly (i.e., display discontinuities) precisely at the November 2 1936 eligibility threshold. To test the validity of this assumption empirically, we conducted a series of baseline balance checks by testing for differences in outcomes at the November 2 1936 date-of-birth threshold. We used the identical analysis approach as for our main outcome analyses except that we used the incidence of the outcome at any time prior to, rather than after, the start of the HZ vaccination program on November 1 2016. We used three sets of outcomes for our baseline balance checks. The first set of outcomes was the 15 most common clinical diagnoses in our data. The second set was indicators of prior uptake of preventive health services that were available in our data. These were uptake of common vaccinations in older age other than for HZ (namely the influenza vaccine, the pneumococcal polysaccharide vaccine, and the diphtheria, pertussis, and tetanus (DPT) vaccine), use of antihypertensive medication, use of statin medications, and participation in colorectal or breast cancer screening (defined, as per Australian cancer screening guidelines,[25,26] as uptake of fecal occult blood testing for colorectal cancer screening and mammography for breast cancer screening). The third set of outcomes were dementia diagnoses prior to November 1 2016 and the prevalence of known risk factors for dementia available in our data. These risk factors were obesity, current smoking, hypertension, diabetes, hyperlipidemia, antihypertensive use, and statin use.

*Testing for confounding:*
The key advantage of our RD approach is that a potential confounding variable only biases our analysis if it changes abruptly (i.e., displays a discontinuity) at exactly the November 2 1936 date-of-birth eligibility threshold.[14,15] Such a discontinuity could occur if another intervention also used November 2 1936 as its date-of-birth eligibility criterion and had an effect on dementia incidence. We conducted two types of tests to investigate the possible presence of such a competing intervention. First, we implemented the same RD analysis as for new dementia diagnoses (i.e., our primary analysis) for new diagnoses of each of the 15 most common clinical diagnoses during the follow-up period in our data. If another intervention existed that used the identical date-of-birth eligibility threshold as the HZ vaccination program and was not specific to dementia, then we may expect this intervention to also affect health outcomes other than dementia. Second, we reasoned that if November 2 was used as an annual date-of-birth eligibility threshold by another intervention, then we would expect to see effects on dementia of the November 2 threshold not merely for the birth year 1936, but also for other birth years. We,

thus, conducted the same RD analysis for new dementia diagnoses as we did for the November 2 1936 date-of-birth eligibility threshold (i.e., our primary analysis) for each of the three years prior to and after 1936 (i.e., date-of-birth eligibility thresholds of November 2 1933, November 2 1934, November 2 1935, November 2 1937, November 2 1938, and November 2 1939). In these tests, given that the maximum follow-up period was shorter for more recent birth years, we restricted the follow-up period for each test to four years such that each test had the same length of follow-up period. However, we additionally conducted these tests using the maximum follow-up period (i.e., until March 27 2024) available for each test.

In our design, there are potential sources of selection bias that we want to take into account. In particular, it is possible that practices carry more complete and accurate records on “active” patients than those who do not visit their GP regularly. Relatedly, other data quality issues, like missing patient IDs or dementia diagnosis dates, may also be influenced both by eligibility for a HZ vaccination and dementia diagnoses, such that removing patients with these data issues could introduce a spurious or non-causal relationship. These sample restrictions would only introduce biases if these data quality issues varied at the date-of-birth eligibility threshold. To assuage concerns about selection bias and potential conditioning on colliders, we perform the following tests. First, we check if there is a discontinuity in either “active” status (using the Royal College of Australian General Practitioners’ definition of active status: patients who visit their GP at least three times in the last two years) or data quality issues at the date-of-birth eligibility threshold (**Figure S14**). We find no statistically significant discontinuity in active patient status (-2.7 percentage points, 95% CI: -7.0, 1.5, p = 0.206). We also fail to find that missing dementia diagnosis dates vary at the date-of-birth eligibility threshold (-0.018 percentage points, 95% CI: -0.3, 0.3, p = 0.911). We do, however, find a discontinuity in missing patient IDs at the threshold (-2.9 percentage points, 95% CI: -4.9, -0.8, p = 0.006). However, when we add patients with missing IDs back into the data (and assume that they are unique observations), we still find a protective effect of eligibility for a free HZ vaccination on dementia (-1.8 percentage points, 95% CI: -3.3, -0.2, p = 0.023).

*Robustness checks:*
We conducted additional robustness checks to those described above. First, we implemented our analysis among “active” patients only, reasoning that delays in the diagnosis of dementia among this patient cohort are likely to be less common. Using the definition of the Royal Australian College of General Practitioners (RACGP), we considered those patients as “active” who visited their GP at least three times in the two years preceding the end of our dataset.[27] This group comprised 61.2% (n=61,903 patients) of our primary analysis cohort. Second, we implemented our analysis both when including and when excluding those patients who had received a diagnosis of dementia prior to the start of the HZ vaccination program on November 1 2016 (n = 94). Third, we verified that our findings were robust to different choices of i) grace periods (i.e., time periods since November 1 2016 after which follow-up time was considered to begin to allow for the time needed for a full immune response to develop after vaccine administration), ii) the bandwidth drawn around the November 2 1936 threshold by using bandwidths ranging from one-half to twice the MSE-optimal bandwidth used in our primary analysis, iii) kernel weights by using both uniform and triangular kernel weights, and iv) functional form assumptions by implementing the analysis using both local linear and quadratic polynomial regression.

**Text S3: Regression Equations**

*Regression discontinuity:*
The regression discontinuity design is:

$$Y_i = \alpha + \beta_1 * (WOB_i - c_0) + \beta_2 * D_i + \beta_3 * D_i * (WOB_i - c_0) + \varepsilon_i$$

where Y represents the outcome of patient I, WOB denotes a patient's week of birth, $c_0$ is the date-of-birth eligibility threshold, ($WOB_i - c_0$) denotes the difference (in weeks) between a patient's week of birth and the eligibility threshold, and $D_i$ is a binary indicator for eligibility for a free HZ vaccine. The key coefficient is $\beta_2$, which gives us the discontinuity, or the difference in dementia diagnoses between patients who were eligible and ineligible for a free HZ vaccine, at the date-of-birth eligibility threshold.

In another specification, we implemented the regression discontinuity design using quadratic polynomials to model the assignment variable (i.e., week of birth relative to the date-of-birth eligibility threshold). This specification is:

$$Y_i = \alpha + \beta_1 * (WOB_i - c_0) + \beta_2 * D_i + \beta_3 * (WOB_i - c_0)^2 + \beta_4 * D_i * (WOB_i - c_0) + \beta_5 * D_i * (WOB_i - c_0)^2 + \varepsilon_i$$

As before, the key coefficient is $\beta_2$.

*Treatment effect heterogeneity:*
We examined heterogeneous treatment effects by gender. We did this in two ways. First, we simply subset the data and implemented the analysis among women only and among men only. Second, to more formally test for treatment effect heterogeneity, we used an interaction term to measure changes in the discontinuity across these sub-groups. We used the rdrobust package in R for the first approach, whereas we used OLS to estimate interaction models. The regression equation with interaction terms for gender was:

$$Y_i = \alpha + \beta_1 * (WOB_i - c_0) + \beta_2 * D_i + \beta_3 * FEMALE_i + \beta_4 * D_i * (WOB_i - c_0) + \beta_5 * D_i * FEMALE_i + \beta_6 * (WOB_i - c_0) * FEMALE_i + \beta_7 * D_i * (WOB_i - c_0) * FEMALE_i + \varepsilon_i$$

The interaction term D*(WOB-$c_0$) allowed for the slope of the regression line to differ on either side of the date-of-birth eligibility threshold. Adding the terms (WOB-$c_0$)*FEMALE and D*(WOB-$c_0$)*FEMALE allowed the slopes to vary by gender. The parameter $\beta_5$ identified the gender difference in the effect of eligibility for HZ vaccination on the outcome.

*Comparative regression discontinuity:*
We used an alternative methodological approach, called the comparative regression discontinuity (CRD) design, as a secondary analysis approach in this study. Comparative regression discontinuity leverages an additional source of untreated data. In our case, these data were dementia diagnoses from older patients in our sample that were never eligible for a free HZ vaccine. The model for this approach was:

$$Y_i = \alpha + \beta_1 * (WOB_i - c_0) + \beta_2 * P_i + \beta_3 * P_i * D_i + \beta_4 * P_i * D_i * (WOB_i - c_0) + \varepsilon_i$$

where Y represented the outcome, WOB denoted a patient's week of birth, $c_0$ was the date-of-birth eligibility threshold, $P_i$ was an indicator for the main cohort, and $D_i$ indicated whether the patient was eligible for a free vaccine after November 1, 2016. The coefficient of interest here was $\beta_3$, which provided the abrupt change in the outcome at the date-of-birth eligibility threshold.

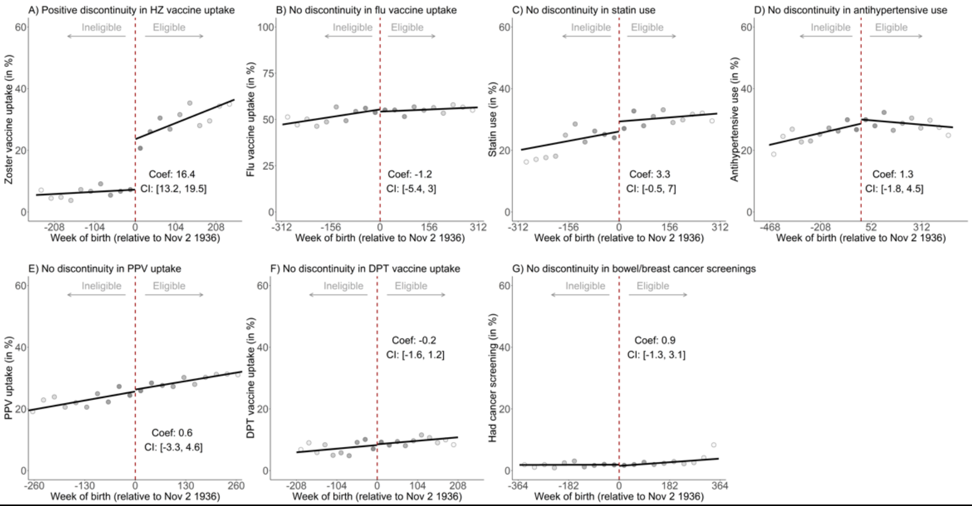


| Sample sizes on ineligible side of the cutoff | | | | | | | | | | |
|---|---|---|---|---|---|---|---|---|---|---|
| **Bin number** | 1 | 2 | 3 | 4 | 5 | 6 | 7 | 8 | 9 | 10 |
| HZ vaccination | 254 | 246 | 250 | 293 | 372 | 313 | 340 | 349 | 417 | 386 |
| Influenza vaccination | 253 | 257 | 297 | 315 | 343 | 428 | 393 | 414 | 492 | 461 |
| Statin use | 271 | 258 | 289 | 304 | 329 | 438 | 383 | 404 | 473 | 461 |
| Antihypertensive use | 219 | 261 | 324 | 365 | 408 | 472 | 529 | 648 | 652 | 741 |
| PPV | 230 | 271 | 272 | 311 | 309 | 389 | 369 | 391 | 440 | 418 |
| DPT vaccination | 205 | 233 | 239 | 299 | 262 | 277 | 270 | 317 | 347 | 298 |
| Cancer screening | 262 | 277 | 309 | 337 | 360 | 422 | 497 | 476 | 541 | 544 |

| Sample sizes on eligible side of the cutoff | | | | | | | | | | |
|---|---|---|---|---|---|---|---|---|---|---|
| **Bin number** | 1 | 2 | 3 | 4 | 5 | 6 | 7 | 8 | 9 | 10 |
| HZ vaccination | 440 | 534 | 489 | 510 | 548 | 580 | 692 | 623 | 652 | 709 |
| Influenza vaccination | 539 | 644 | 593 | 662 | 716 | 803 | 773 | 801 | 898 | 910 |
| Statin use | 539 | 614 | 597 | 623 | 682 | 803 | 746 | 770 | 829 | 931 |
| Antihypertensive use | 902 | 916 | 1,053 | 1,174 | 1,203 | 1,323 | 1,424 | 1,673 | 1,702 | 2,007 |
| PPV | 479 | 581 | 536 | 587 | 606 | 694 | 701 | 699 | 735 | 811 |
| DPT vaccination | 347 | 424 | 436 | 372 | 435 | 442 | 487 | 520 | 521 | 525 |
| Cancer screening | 651 | 712 | 746 | 808 | 895 | 892 | 948 | 1,040 | 1,055 | 1,191 |

**Fig. S1:** The date-of-birth eligibility cutoff led to a large discontinuity in HZ vaccination receipt but there was no such jump at the cutoff in the uptake of other preventive interventions.[1,2,3]

Abbreviations: HZ = Herpes Zoster; PPV = Pneumococcal Polysaccharide Vaccine; DPT = Diphtheria, Tetanus, and Pertussis

[1] Cancer screening referred to the uptake of colorectal or breast cancer screening. As per Australian cancer screening guidelines, this was defined as uptake of fecal occult blood testing (for colorectal cancer screening) and mammography (for breast cancer screening).[25,26]

[2] Uptake of preventive health services were measured during our follow-up period of November 1 2016 (the start date of the HZ vaccination program) to March 27 2024.

[3] Grey dots in panel A show the mean value for each 26-week increment in week of birth. In panels B through G, the increments were 31, 30, 47, 28, 21, and 36 weeks, respectively. Increments were derived by dividing the mean squared error-optimal bandwidth for each analysis by 10. The shading of the dots is in proportion to the weight that observations from each increment received in the analysis.

| | Eligible patients at the threshold | Ineligible patients at the threshold | Discontinuity at the threshold (95% CI) | P value |
|---|---|---|---|---|
| Hypertension | 17.9 | 18.0 | -0.2 (-3.4, 3.1) | 0.92 |
| Hyperlipidemia | 11.3 | 11.7 | -0.3 (-2.8, 2.1) | 0.79 |
| Respiratory conditions | 6.0 | 7.2 | -1.2 (-3.5, 1.1) | 0.30 |
| Nonhematologic cancers | 10.2 | 9.7 | 0.5 (-1.7, 2.8) | 0.64 |
| Osteoarthritis | 10.2 | 9.1 | 1.1 (-1.7, 3.9) | 0.43 |
| Heart disease | 10.6 | 9.5 | 1.0 (-1.9, 4.0) | 0.49 |
| Diabetes mellitus | 6.0 | 5.5 | 0.5 (-1.3, 2.3) | 0.55 |
| Depression | 2.0 | 2.1 | -0.2 (-1.3, 1.0) | 0.76 |
| Osteoporosis | 4.8 | 6.6 | -1.9 (-3.8, 0.1) | 0.06 |
| Back pain | 2.7 | 1.9 | 0.8 (-0.3, 1.9) | 0.15 |
| Gout | 2.8 | 2.9 | -0.1 (-1.4, 1.2) | 0.84 |
| Stroke or TIA | 2.8 | 2.6 | 0.2 (-1.2, 1.6) | 0.79 |
| Hematologic conditions | 2.0 | 1.6 | 0.3 (-0.7, 1.4) | 0.51 |
| Chronic kidney disease | 1.9 | 1.7 | 0.2 (-0.8, 1.2) | 0.68 |
| Influenza vaccination | 26.9 | 26.8 | 0.1 (-3.8, 4.0) | 0.96 |
| PPV | 20.3 | 19.3 | 0.9 (-2.6, 4.5) | 0.61 |
| Statin use | 8.2 | 6.9 | 1.3 (-0.9, 3.4) | 0.25 |
| Antihypertensive use | 7.2 | 5.2 | 2.1 (-0.2, 4.3) | 0.07 |
| DPT vaccination | 4.4 | 5.2 | -0.8 (-2.5, 0.8) | 0.33 |
| Cancer screening | 0.8 | 1.5 | -0.7 (-1.8, 0.4) | 0.19 |

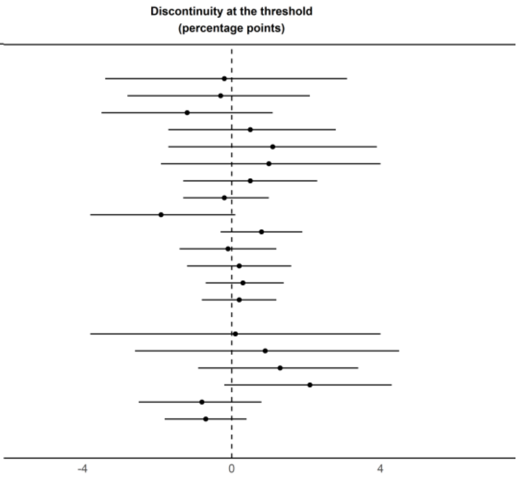


**Fig. S2:** Baseline balance checks using the 15 most common diagnoses in the PenCS data and indicators of preventive health services uptake prior to the start of the vaccination program.[1,2,3,4,5]

Abbreviations: TIA = Transient Ischemic Attack; PPV = Pneumococcal Polysaccharide Vaccine; DPT = Diphtheria, Tetanus, and Pertussis

[1] All diagnoses and uptake of preventive health services were measured prior to the start of the vaccination program (November 1 2016).

[2] Dots show the point estimate and horizontal bars the 95% confidence interval.

[3] The codes used to define each condition are shown in Table S1.

[4] Cancer screening refers to the uptake of colorectal or breast cancer screening. As per Australian cancer screening guidelines, this was defined as uptake of fecal occult blood testing (for colorectal cancer screening) and mammography (for breast cancer screening).[25,26]

[5] Note that COVID-19 is omitted from this analysis of clinical diagnoses because there was no incidence of COVID-19 prior to November 1 2016.

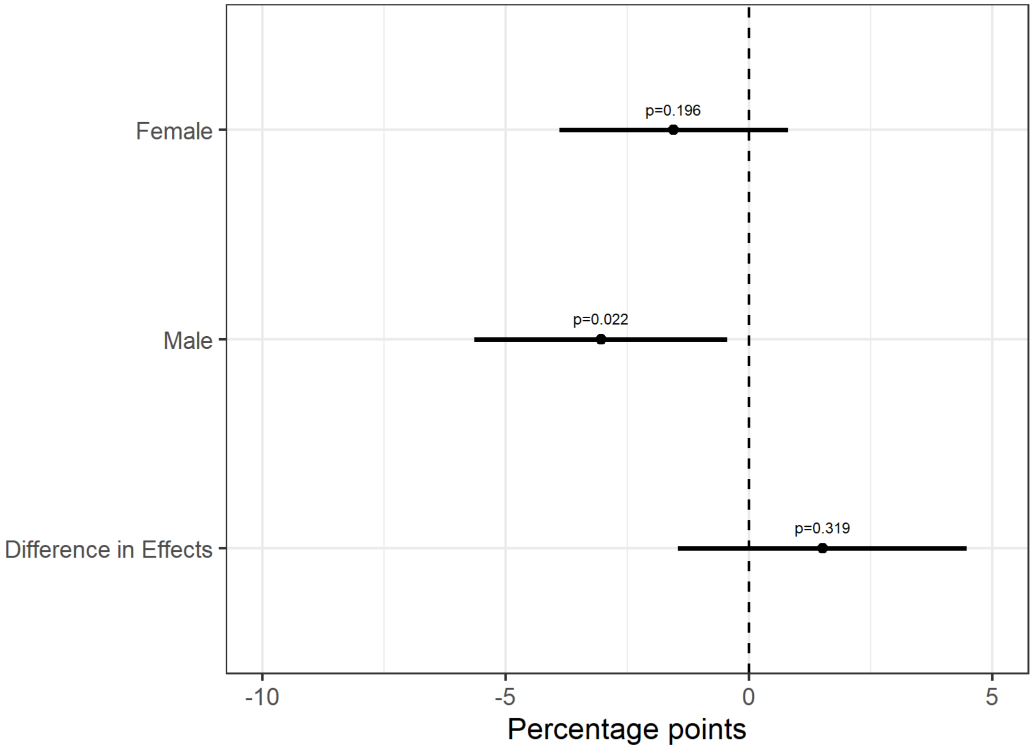


**Fig. S3:** Effect of being eligible for HZ vaccination on new diagnoses of dementia over a 7.4-year follow-up period, by gender.[1,2,3]

[1] “Female” and “Male” depict the point estimates and 95% confidence intervals for the effect of herpes zoster vaccination eligibility on new diagnoses of dementia when the analysis was implemented separately among women and men, respectively.

[2] “Difference in Effects” depicts the difference for (Female – Male) using the interaction model described in Text S1.

[3] Effect estimates were obtained using regression discontinuity with triangular kernel weights, MSE-optimal bandwidths, and a local linear polynomial.

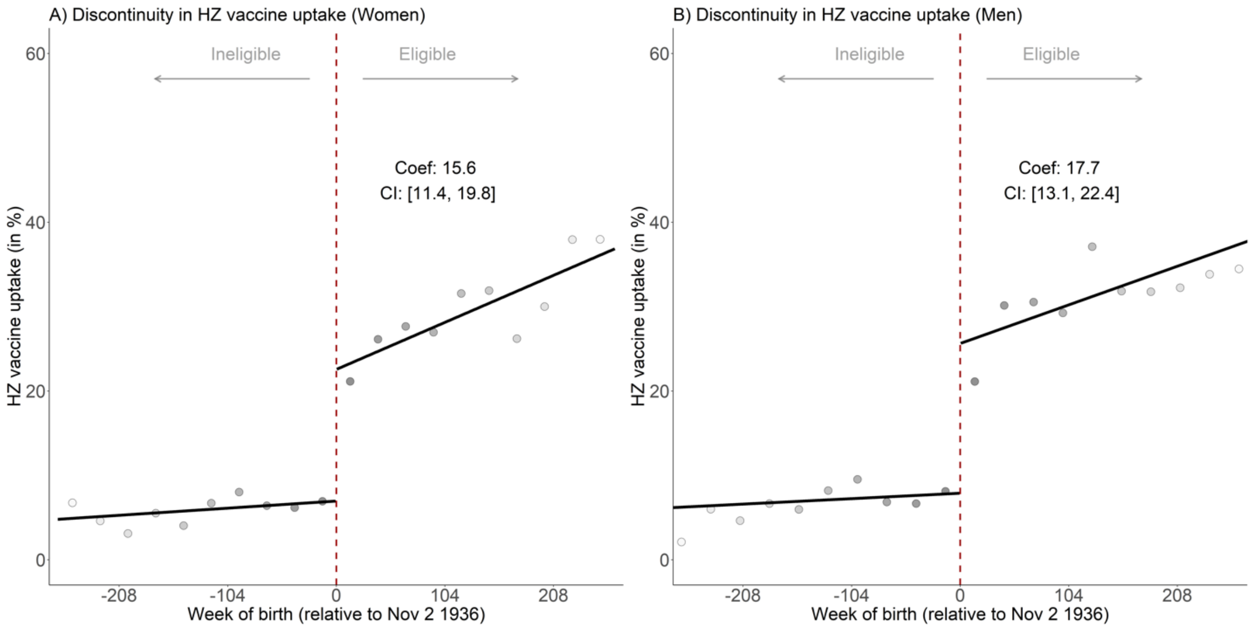


**Fig. S4:** The discontinuity in HZ vaccine uptake was similar between women (panel A) and men (panel B).[1,2]

Abbreviations: HZ = Herpes Zoster

[1] Grey dots show the mean value for each 27-week increment in week of birth in panel A and for each 28-week increment in week of birth in panel B. The shading of the dots is in proportion to the weight that observations in each increment received in the analysis.

[2] Effect estimates were obtained using regression discontinuity with triangular kernel weights, MSE-optimal bandwidths, and a local linear polynomial.

| **Sample sizes on ineligible side of the cutoff** | | | | | | | | | | |
|---|---|---|---|---|---|---|---|---|---|---|
| **Bin number** | 1 | 2 | 3 | 4 | 5 | 6 | 7 | 8 | 9 | 10 |
| Women | 148 | 152 | 161 | 181 | 173 | 194 | 212 | 203 | 243 | 231 |
| Men | 95 | 100 | 108 | 105 | 134 | 195 | 147 | 161 | 180 | 185 |

| **Sample sizes on eligible side of the cutoff** | | | | | | | | | | |
|---|---|---|---|---|---|---|---|---|---|---|
| **Bin number** | 1 | 2 | 3 | 4 | 5 | 6 | 7 | 8 | 9 | 10 |
| Women | 246 | 287 | 264 | 293 | 282 | 373 | 351 | 350 | 369 | 424 |
| Men | 232 | 282 | 239 | 270 | 283 | 311 | 340 | 304 | 331 | 380 |

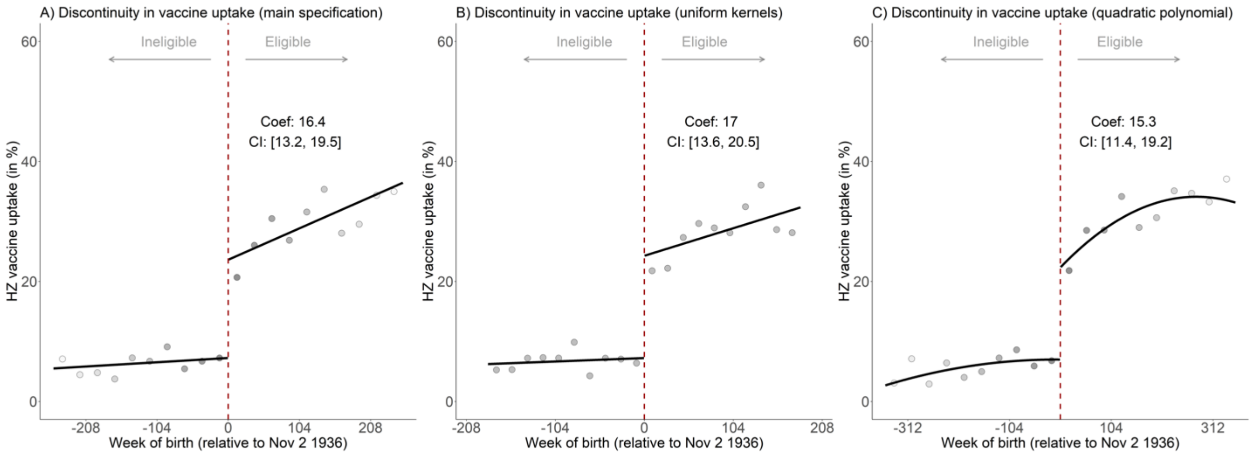


**Fig. S5:** The effect of being eligible for HZ vaccination on HZ vaccination uptake using different kernel weights and functional forms.[1,2,3,4]

Abbreviations: HZ = Herpes Zoster

[1] Panel A presents our primary specification (which is also shown in Figure S1), which utilized a linear polynomial, triangular kernel weights, and the MSE-optimal bandwidth.

[2] Panel B used uniform instead of triangular kernel weights.

[3] Panel C used a quadratic polynomial to model the running variable (i.e., week of birth relative to November 2, 1936) instead of a linear polynomial.

[4] Grey dots in panel A show the mean value for each 26-week increment in week of birth. In panels B and C, the increments were 18 and 36 weeks, respectively. Increments were derived by dividing the MSE-optimal bandwidth for each analysis by 10. The shading of the dots is in proportion to the weight that observations from each increment received in the analysis.

| **Sample sizes on ineligible side of the cutoff** | | | | | | | | | | |
|---|---|---|---|---|---|---|---|---|---|---|
| **Bin number** | 1 | 2 | 3 | 4 | 5 | 6 | 7 | 8 | 9 | 10 |
| Main specification | 254 | 246 | 250 | 293 | 372 | 313 | 340 | 349 | 417 | 386 |
| Uniform kernels | 209 | 207 | 277 | 219 | 235 | 253 | 257 | 277 | 283 | 281 |
| Quadratic polynomial | 259 | 281 | 308 | 342 | 348 | 422 | 497 | 476 | 541 | 544 |

| **Sample sizes on eligible side of the cutoff** | | | | | | | | | | |
|---|---|---|---|---|---|---|---|---|---|---|
| **Bin number** | 1 | 2 | 3 | 4 | 5 | 6 | 7 | 8 | 9 | 10 |
| Main specification | 440 | 534 | 489 | 510 | 548 | 580 | 692 | 623 | 652 | 709 |
| Uniform kernels | 303 | 374 | 366 | 344 | 363 | 395 | 376 | 441 | 412 | 512 |
| Quadratic polynomial | 651 | 712 | 746 | 808 | 876 | 885 | 940 | 1,038 | 1,046 | 1,190 |

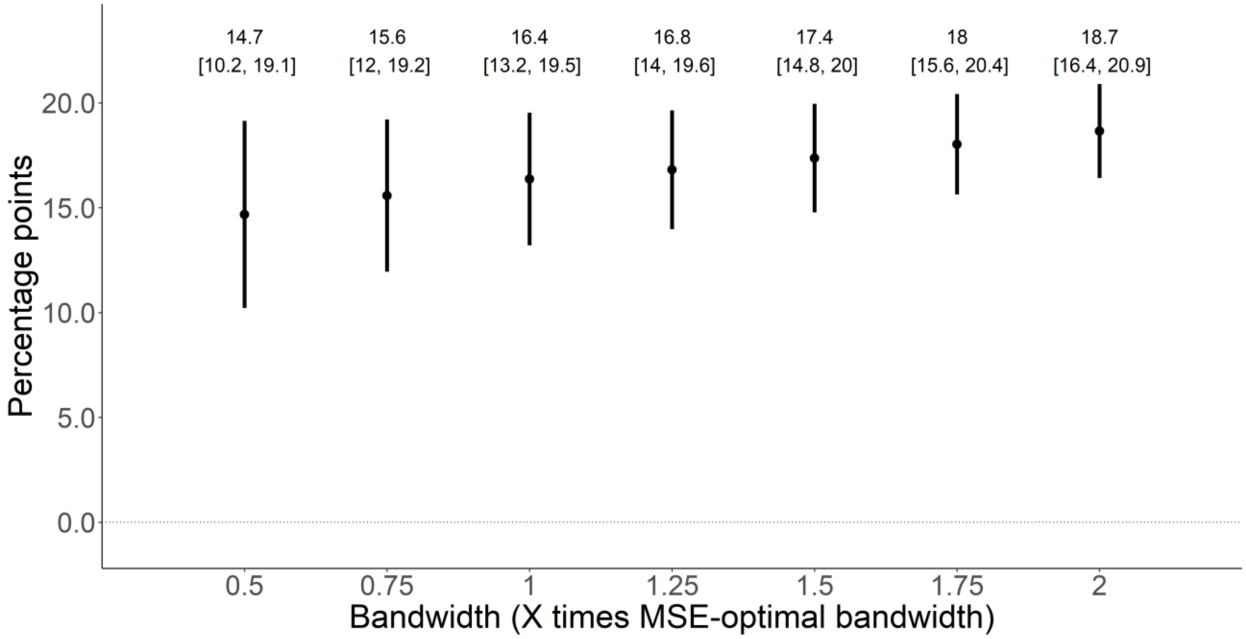


**Fig. S6:** The effect of being eligible for HZ vaccination on HZ vaccination uptake using different bandwidths.[1,2]

Abbreviations: MSE = Mean Squared Error

[1] Vertical bars depict 95% confidence intervals.

[2] All specifications used triangular kernels and a local linear polynomial to model week of birth relative to the date-of-birth eligibility threshold.

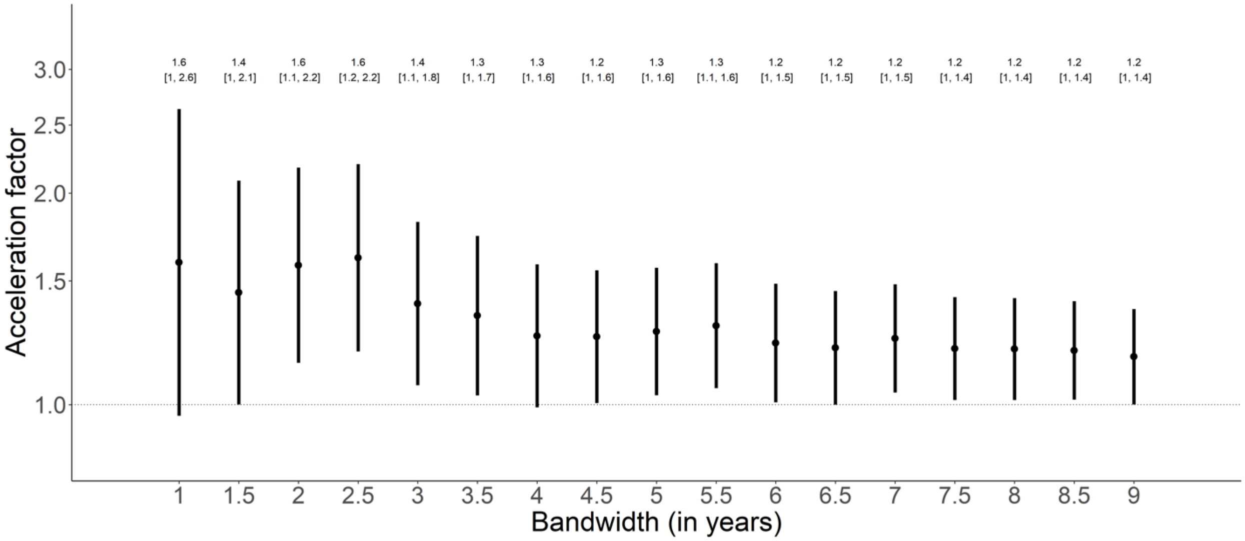


**Fig. S7:** The acceleration factor across different bandwidths, which indicates how much longer patients eligible for HZ vaccination take to be diagnosed with dementia than ineligible patients.[1,2]

[1] Vertical bars depict 95% confidence intervals.

[2] The y-axis is logarithmically transformed.

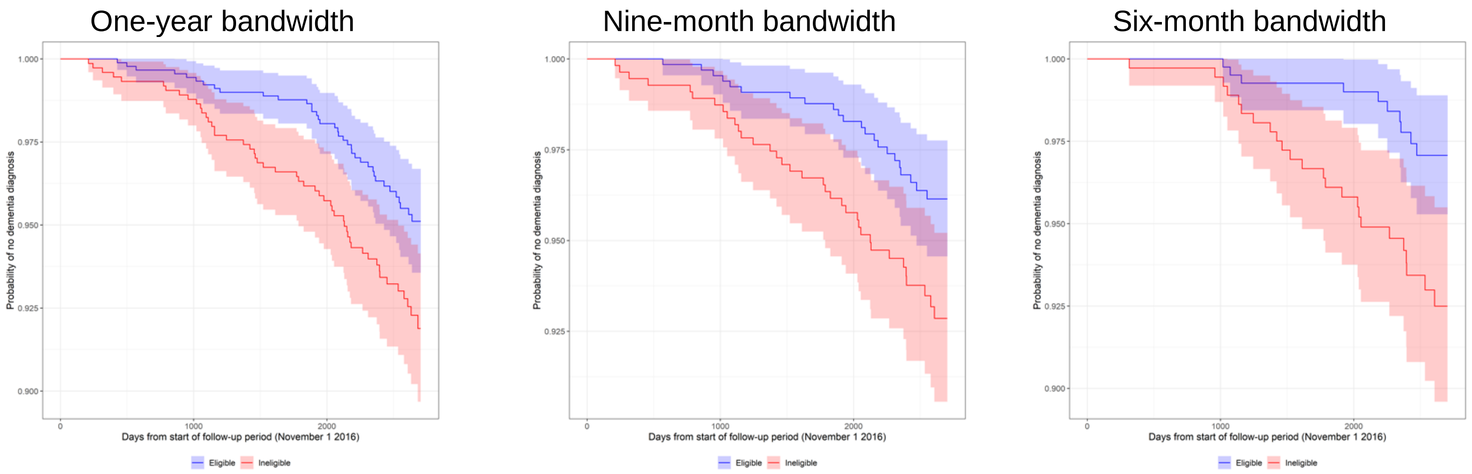


**Fig. S8:** Survival functions (Kaplan-Meier plots) for patients eligible and ineligible for HZ vaccination.[1]

[1] Each graph shows the survival functions for eligible patients in blue and ineligible patients in red, between the start of the immunization program on November 1 2016 and the end of the follow-up period (March 27 2024).

One-year bandwidth

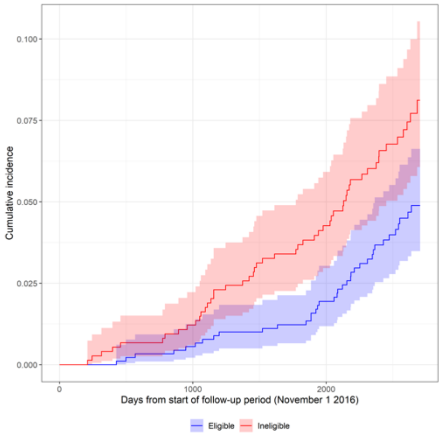


| Characteristic | N | Day 1000 | Day 2000 | p-value[†] |
|---|---|---|---|---|
| Eligibility for free HZ vaccine | 1,818 | | | 0.010 |
| Eligible | | 0.56% (0.21%, 1.2%) | 1.9% (1.2%, 3.0%) | |
| Ineligible | | 1.2% (0.60%, 2.2%) | 4.3% (3.0%, 5.9%) | |

[†] Gray's Test

Nine-month bandwidth

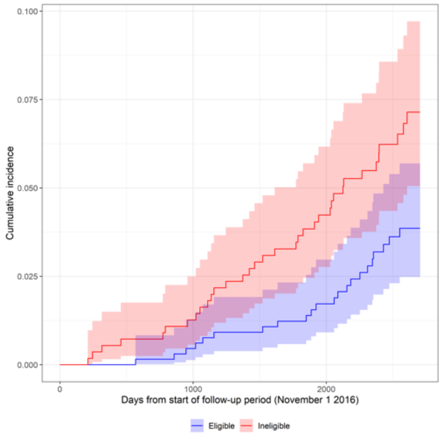


| Characteristic | N | Day 1000 | Day 2000 | p-value[†] |
|---|---|---|---|---|
| Eligibility for free HZ vaccine | 1,341 | | | 0.013 |
| Eligible | | 0.46% (0.13%, 1.3%) | 1.7% (0.91%, 3.0%) | |
| Ineligible | | 1.3% (0.57%, 2.5%) | 4.2% (2.8%, 6.2%) | |

[†] Gray's Test

Six-month bandwidth

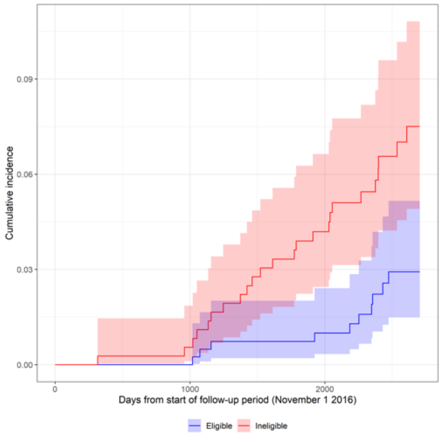


| Characteristic | N | Day 1000 | Day 2000 | p-value[†] |
|---|---|---|---|---|
| Eligibility for free HZ vaccine | 856 | | | 0.005 |
| Eligible | | 0.00% (—%, —%) | 1.0% (0.34%, 2.4%) | |
| Ineligible | | 0.55% (0.11%, 1.9%) | 4.2% (2.4%, 6.6%) | |

[†] Gray's Test

**Fig. S9:** Cumulative incidence curves for patients eligible and ineligible for HZ vaccination.[1,2]

Abbreviations: HZ = Herpes Zoster

[1] Each graph shows the cumulative incidence curves for eligible patients in blue and ineligible patients in red, between the start of the immunization program on November 1 2016 and the end of the follow-up period (March 27 2024).

[2] Gray's test is performed for each bandwidth and presented below the corresponding graph.

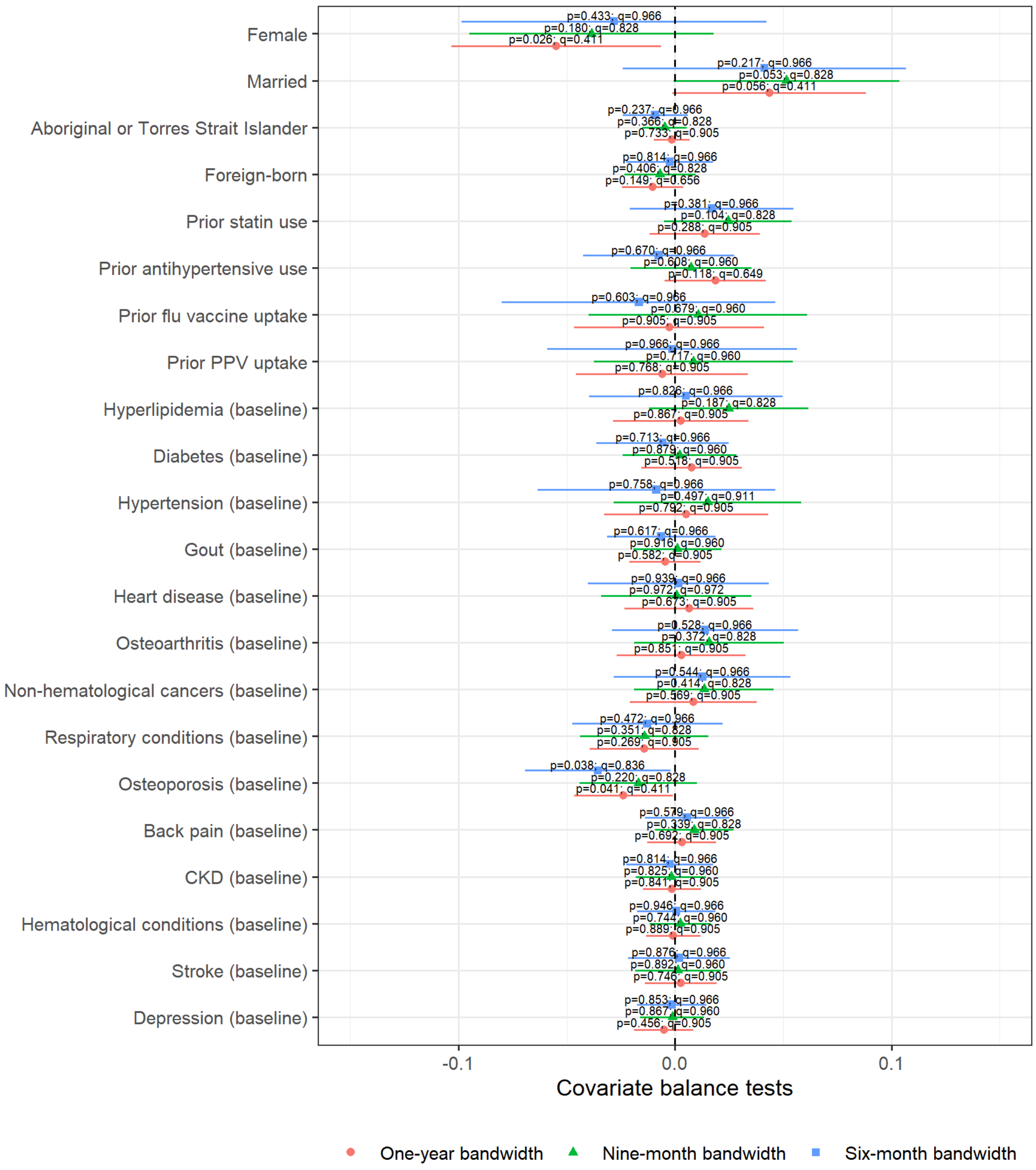


**Fig. S10:** Differences in covariates between patients eligible and ineligible for HZ vaccination, for one-year, nine-month, and six-month bandwidths.[1,2]

Abbreviations: PPV = Pneumococcal Polysaccharide Vaccine, CKD = Chronic Kidney Disease

[1] Each row shows the result of a two-sample t-test comparing eligible and ineligible patients.

[2] Dots indicate difference in means, and horizontal bars depict 95% confidence intervals.

[3] We present sharpened q-values (alongside p-values) that represent the false positive rate.[28]

| | Effect size, (95% CI) | p-value |
|---|---|---|
| Main specification | -1.5 (-2.7, -0.2) | 0.020 |
| Triangular kernels | -1.8 (-3.2, -0.4) | 0.012 |
| Polynomial order = 2 | -2.1 (-3.5, -0.7) | 0.003 |
| 3 | -2.9 (-4.3, -1.5) | < 0.001 |
| 4 | -3.3 (-4.7, -1.9) | < 0.001 |
| Bandwidth = 9 years | -1.5 (-2.7, -0.2) | 0.022 |
| 8 years | -1.6 (-2.9, -0.3) | 0.016 |
| 7 years | -1.7 (-3.0, -0.3) | 0.016 |
| 6 years | -1.3 (-2.7, 0.2) | 0.081 |
| 5 years | -1.8 (-3.3, -0.2) | 0.026 |
| 4 years | -1.7 (-3.4, -0.1) | 0.043 |
| 3 years | -2.1 (-4.0, -0.1) | 0.036 |
| 2 years | -3.5 (-5.8, -1.2) | 0.003 |
| 1 year | -4.3 (-7.4, -1.2) | 0.006 |
| Active patients | -2.3 (-4.2, -0.5) | 0.014 |
| Excl. patients w/baseline dementia | -1.5 (-2.7, -0.3) | 0.017 |

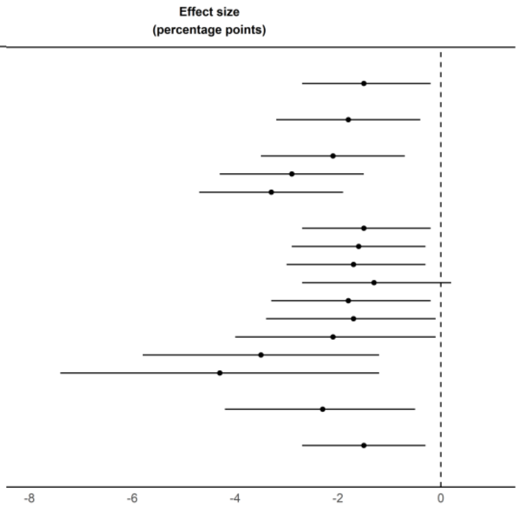


**Fig. S11:** Effect of being eligible for HZ vaccination on new diagnoses of dementia over a 7.4-year follow-up period, using the comparative regression discontinuity design.[1,2]

Abbreviations: HZ = Herpes Zoster

[1] The main specification used uniform kernel weights, and a linear polynomial for the running variable (i.e., week of birth relative to November 2, 1936).

[2] All specifications used a bandwidth of 482 weeks for both the main and comparison cohorts, unless otherwise stated.

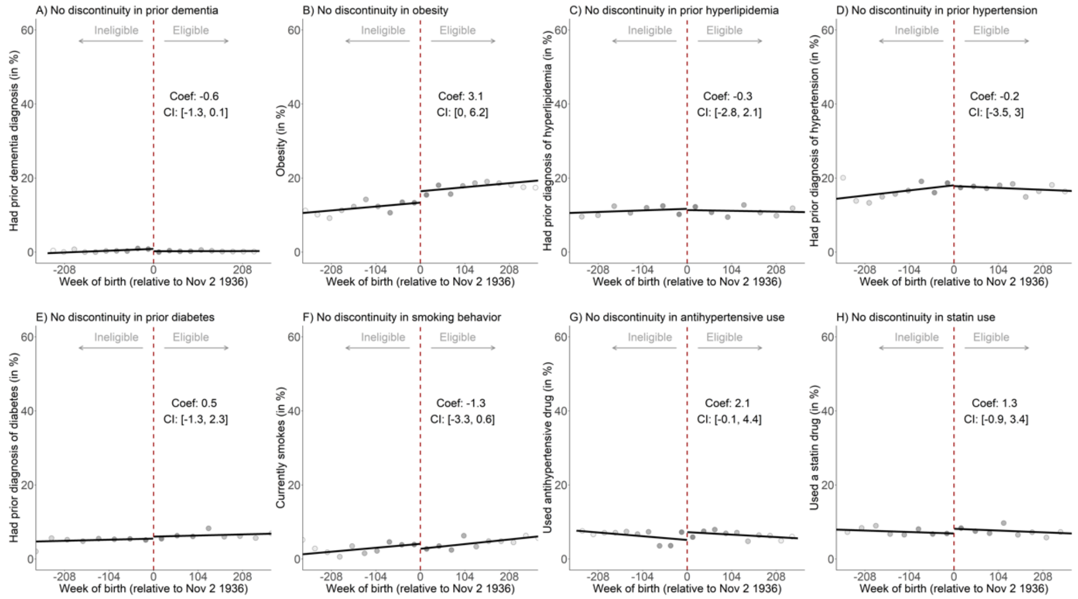


**Fig. S12:** There was balance at baseline across the November 2 1936 date-of-birth threshold in dementia risk factors and dementia diagnoses recorded prior to the start of the HZ vaccination program.[1,2]

[1] Each outcome variable was defined as having been recorded at any time prior to November 1 2016.

[2] Grey dots in panel A show the mean value for each 25-week increment in week of birth. In panels B through H, the increments were 28, 38, 31, 37, 29, 26, and 33 weeks, respectively. Increments were derived by dividing the MSE-optimal bandwidth for each analysis by 10. The shading of the dots is in proportion to the weight that observations from each increment received in the analysis.

| **Sample sizes on ineligible side of the cutoff** | | | | | | | | | | |
|---|---|---|---|---|---|---|---|---|---|---|
| **Bin number** | 1 | 2 | 3 | 4 | 5 | 6 | 7 | 8 | 9 | 10 |
| Baseline dementia | 244 | 248 | 264 | 264 | 363 | 319 | 313 | 361 | 404 | 363 |
| Obesity | 241 | 267 | 283 | 301 | 317 | 401 | 381 | 386 | 438 | 435 |
| Hyperlipidemia | 253 | 270 | 324 | 345 | 382 | 411 | 519 | 501 | 561 | 578 |
| Hypertension | 266 | 254 | 289 | 308 | 341 | 427 | 391 | 398 | 492 | 461 |
| Diabetes | 259 | 278 | 294 | 355 | 365 | 415 | 491 | 488 | 552 | 564 |
| Smoking | 135 | 142 | 164 | 166 | 201 | 258 | 230 | 239 | 287 | 308 |
| Antihypertensive use | 247 | 261 | 259 | 284 | 353 | 338 | 342 | 362 | 416 | 403 |
| Statin use | 252 | 270 | 303 | 322 | 356 | 416 | 443 | 445 | 504 | 508 |

| **Sample sizes on eligible side of the cutoff** | | | | | | | | | | |
|---|---|---|---|---|---|---|---|---|---|---|
| **Bin number** | 1 | 2 | 3 | 4 | 5 | 6 | 7 | 8 | 9 | 10 |
| Baseline dementia | 423 | 525 | 483 | 473 | 534 | 576 | 637 | 614 | 618 | 648 |
| Obesity | 505 | 576 | 536 | 615 | 639 | 699 | 715 | 717 | 730 | 839 |
| Hyperlipidemia | 695 | 754 | 783 | 841 | 972 | 977 | 1,046 | 1,101 | 1,153 | 1,386 |
| Hypertension | 539 | 614 | 610 | 632 | 706 | 785 | 760 | 772 | 867 | 938 |
| Diabetes | 677 | 732 | 756 | 810 | 933 | 937 | 970 | 1,055 | 1,093 | 1,253 |
| Smoking | 331 | 402 | 369 | 429 | 479 | 478 | 498 | 489 | 603 | 603 |
| Antihypertensive use | 457 | 551 | 493 | 538 | 554 | 607 | 686 | 647 | 661 | 768 |
| Statin use | 600 | 677 | 648 | 710 | 828 | 845 | 828 | 932 | 959 | 1,004 |

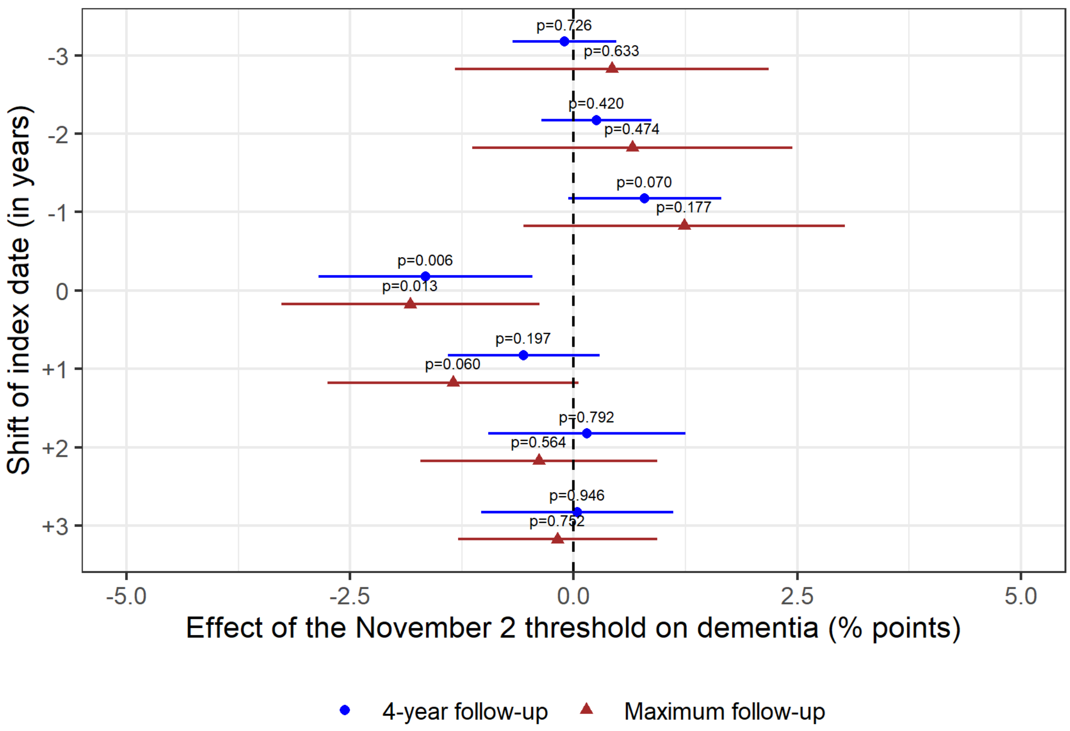


**Fig. S13:** The November 2 date-of-birth threshold only had a protective effect for the occurrence of dementia in the birth year (1936) that was used by the HZ vaccination program as eligibility criterion.[1,2]

Abbreviations: HZ = Herpes Zoster

[1] -3 refers to 1933, -2 to 1934, -1 to 1935, 0 to 1936, +1 to 1937, +2 to 1938, and +3 to 1939.

[2] For each of the shifts, we imposed a follow-up period of 4 years, since that is the maximum number of full years available for the most recent threshold (i.e., November 1 1939). As an example, when shifting the index date to November 2 1934, we started the follow-up period on November 1 2014 and the follow-up period ended on November 1 2018. We also show results for the maximum possible follow-up period for each threshold (i.e., ending on March 27, 2024).

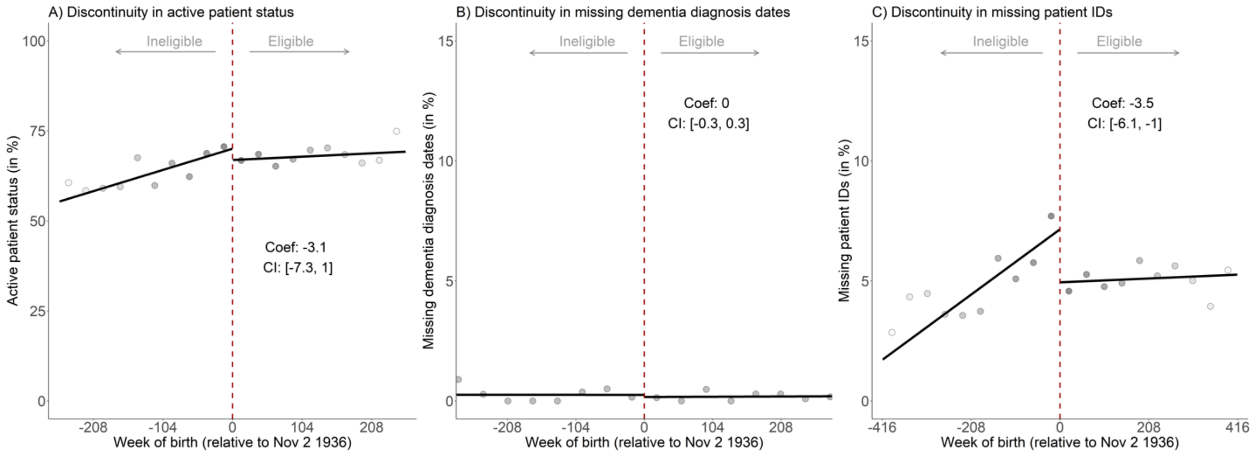


**Fig. S14:** There was balance at baseline across the November 2 1936 date-of-birth threshold in active patient status and missing dementia diagnosis dates, but not for missing patient IDs.[1,2]

[1] These analyses used the full dataset (n = 106,920) without any patients excluded.

[2] Main results were substantively the same when patients with missing patient IDs are included in the analysis.

| **Sample sizes on ineligible side of the cutoff** | | | | | | | | | | |
|---|---|---|---|---|---|---|---|---|---|---|
| **Bin number** | 1 | 2 | 3 | 4 | 5 | 6 | 7 | 8 | 9 | 10 |
| Active patient status | 259 | 267 | 264 | 311 | 373 | 346 | 362 | 390 | 445 | 422 |
| Missing dementia diagnosis date | 262 | 292 | 335 | 355 | 393 | 443 | 530 | 533 | 595 | 630 |
| Missing patient IDs | 245 | 300 | 335 | 360 | 421 | 455 | 572 | 570 | 625 | 701 |

| **Sample sizes on eligible side of the cutoff** | | | | | | | | | | |
|---|---|---|---|---|---|---|---|---|---|---|
| **Bin number** | 1 | 2 | 3 | 4 | 5 | 6 | 7 | 8 | 9 | 10 |
| Active patient status | 464 | 572 | 523 | 563 | 557 | 643 | 719 | 699 | 685 | 785 |
| Missing dementia diagnosis date | 731 | 794 | 826 | 868 | 1,029 | 1,032 | 1,063 | 1,165 | 1,215 | 1,440 |
| Missing patient IDs | 809 | 854 | 924 | 1,059 | 1,094 | 1,132 | 1,261 | 1,316 | 1,497 | 1,615 |
